# Exome-wide analysis of congenital kidney anomalies reveals new genes and shared architecture with developmental disorders

**DOI:** 10.1101/2024.11.05.24316672

**Authors:** Hila Milo Rasouly, Sarath Babu Krishna Murthy, Natalie Vena, Gundula Povysil, Andrew Beenken, Miguel Verbitsky, Shirlee Shril, Iris Lekkerkerker, Atlas Khan, David Fasel, Janewit Wongboonsin, Jeremiah Martino, Juntao Ke, Naama Elefant, Nikita Tomar, Ofek Harnof, Sandy Yang, Sergey Kisselev, Shiraz Bheda, Sivan Reytan-Miron, Tze Y Lim, Anna Jamry-Dziurla, Francesca Lugani, Jun Y Zhang, Maddalena Marasa, Victoria Kolupaeva, Emily E. Groopman, Gina Jin, Iman Ghavami, Kelsey O. Stevens, Arielle C. Coughlin, Byum Hee Kil, Debanjana Chatterjee, Drew Bradbury, Jason Zheng, Karla Mehl, Maria Morban, Rachel Reingold, Stacy Piva, Xueru Mu, Adele Mittrori, Agnieszka Szmigielska, Aleksandra Gliwińska, Andrea Ranghino, Andrew S Bomback, Andrzej Badenski, Anna Latos-Bielenska, Anna Materna-Kiryluk, Antonio Amoroso, Claudia Izzi, Claudio La Scola, David Jonathan Cohen, Domenico Santoro, Dorota Drozdz, Enrico Fiaccadori, Fangming Lin, Francesco Scolari, Francesco Tondolo, Gaetano La Manna, Gerald B Appel, Gian Marco Ghiggeri, Gianluigi Zaza, Giovanni Montini, Giuseppe Masnata, Grażyna Krzemien, Isabella Pisani, Jai Radhakrishnan, Katarzyna Zachwieja, Lauren Monaco, Loreto Gesualdo, Luigi Biancone, Luisa Murer, Malgorzata Mizerska-Wasiak, Marcin Tkaczyk, Marcin Zaniew, Maria K. Borszewska-Kornacka, Maria Szczepanska, Marijan Saraga, Maya K Rao, Monica Bodria, Monika Miklaszewska, Natalie S Uy, Olga Baraldi, Omar Bjanid, Pasquale Esposito, Pasquale Zamboli, Pierluigi Marzuillo, Pietro A Canetta, Przemyslaw Sikora, Rik Westland, Russell J Crew, Shumyle Alam, Stefano Guarino, Susanna Negrisolo, Thomas Hays, Valeria Grandinetti, Velibor Tasic, Vladimir J. Lozanovski, Yasar Caliskan, David Goldstein, Richard P Lifton, Iuliana Ionita-Laza, Krzysztof Kiryluk, Albertien van Eerde, Friedhelm Hildebrandt, Simone Sanna-Cherchi, Ali G Gharavi

## Abstract

Kidney anomalies (KA) are developmental disorders that commonly cause pediatric chronic kidney disease and mortality. We examined rare coding variants in 248 KA trios and 1,742 singleton KA cases and compared them to 22,258 controls. Diagnostic and candidate diagnostic variants were detected in 14.1% of cases. We detected a significant enrichment of rare damaging variants in constrained genes expressed during kidney development and in genes associated with other developmental disorders, suggesting phenotype expansion. Consistent with these data, 18% of KA patients with diagnostic variants had neurodevelopmental or cardiac phenotypes. Extrarenal developmental phenotypes were associated with a higher burden of rare variants. Statistical analyses identified 40 novel candidate genes, 2 of which were confirmed as new KA genes: *ARID3A* and *NR6A1.* This study suggests that many yet-unidentified syndromes would be discoverable with larger cohorts and cross-phenotype analysis, leading to clarification of the genetic and phenotypic spectrum of developmental disorders.

## Introduction

Congenital anomalies of the kidney and urinary tract (CAKUT) are diagnosed in 0.5% of live births ^1,2^ and account for 50% of pediatric kidney failure. CAKUT includes a spectrum of developmental defects affecting the kidney number, size, morphology, and position and the lower urinary tract (outflow abnormalities).^3^ Kidney anomalies (KA) affecting kidney size and morphology (hypoplastic kidneys, dysplastic kidneys, multicystic dysplastic kidneys) and number (kidney agenesis) have a high risk of progression to kidney failure.^3^ Identification of a genetic cause for KA can impact prognosis and clinical management.^4,5^ Although multiple causative genes for KA have been identified, surveys based on modest-sized cohorts indicate that they only account for 5-20% of cases.^6^ The two genes accounting for the highest proportion of genetic diagnoses of KA are *HNF1β* and *PAX2*.^7,8^ Despite the high diagnostic yield, genetic testing is not uniformly implemented in pediatric practice.

While KA most often occur in isolation, they can also manifest in conjunction with developmental defects in other organs. For example, 30% of infants with congenital heart disease (CHD) also have urinary tract defects.^9,10^ Shared pathogenesis of KA and neurodevelopmental disorders, such as autism spectrum disorder (ASD) and intellectual disability (ID), has also been suggested.^6,11^ For example, Bardet-Biedl, CHARGE, Cornelia de Lange syndrome, and Smith-Magenis syndromes are associated with CAKUT, CHD, and ASD. Similarly, structural variants such as the 22q11.2 deletion (DiGeorge syndrome), 7q11.23 deletion (Williams syndrome), 16p11.2 deletions/duplications, and 17q12 deletion are associated with all these disorders. ^12^ However, the possibility of shared genetic architecture between KA and other developmental disorders has not been systematically examined.

The majority of KA is detected in patients without a known family history of disease, suggesting the possibility of *de-novo* variants or recessive disease. The analysis of *de-novo* variants has been a successful approach to novel gene discovery for developmental disorders.^13–16^ Another successful approach is gene-burden case-control association studies, enabling the identification of new genes for schizophrenia, amyotrophic lateral sclerosis^17,18^, retinal diseases^19^, epilepsy^20^, and KA.^21^ Both approaches take advantage of large population databases to restrict the analysis and test enrichment for exceedingly rare, predicted deleterious variants, which are more likely to be associated with developmental anomalies like KA. We hypothesized that similar analyses could identify novel KA-causing genes.

## Results

### High genetic heterogeneity of Kidney Anomalies

The study included a total of 1,990 unrelated probands with KA (defined as unilateral or bilateral renal agenesis, hypoplastic kidney disease, dysplastic kidney disease or multicystic dysplastic kidney disease (**Table 1**) and 518 parents (507 unaffected and 11 affected), as well as 22,258 controls (**Table S1**). For the diagnostic analysis, genes associated with multiple KA Human Phenotype Ontology (HPO) terms, or multiple independent associations with KA in the literature were defined as “known genes” (N=208, **Table S2**), whereas genes with single reports, or limited evidence for causality in the literature were defined as “probable genes” (N=297, **Table S2**). Exome sequencing and microarray analysis identified diagnostic variants in 205 (10.3%) probands across 56 known genes and 25 known structural variants (**Fig. 1a**). Of those, 12 probands had dual diagnoses. In addition, 34 (1.7%) probands harbored candidate diagnostic variants, defined as borderline variants of uncertain significance in known genes (“VUS-high”)^22^, and 41 (2.1%) probands with P/LP variants in genes with emerging associations with KA (“probable genes”, **Fig. 1a, Table S3**). Of the 133 probands with a P/LP variant in a known gene, 52 (39%) had variants in *HNF1β, PAX2, EYA1, DSTYK* or *GREB1L* and 31 (23.3%) were singleton diagnoses (**Fig. 1b**). Of the 72 probands with diagnostic structural variants, 29 (40.3%) had the 17q12 (RCAD) deletion and 8 (11.1%) had single gene deletions identified by exome sequencing (**Fig. 1c**).

**Figure 1:**
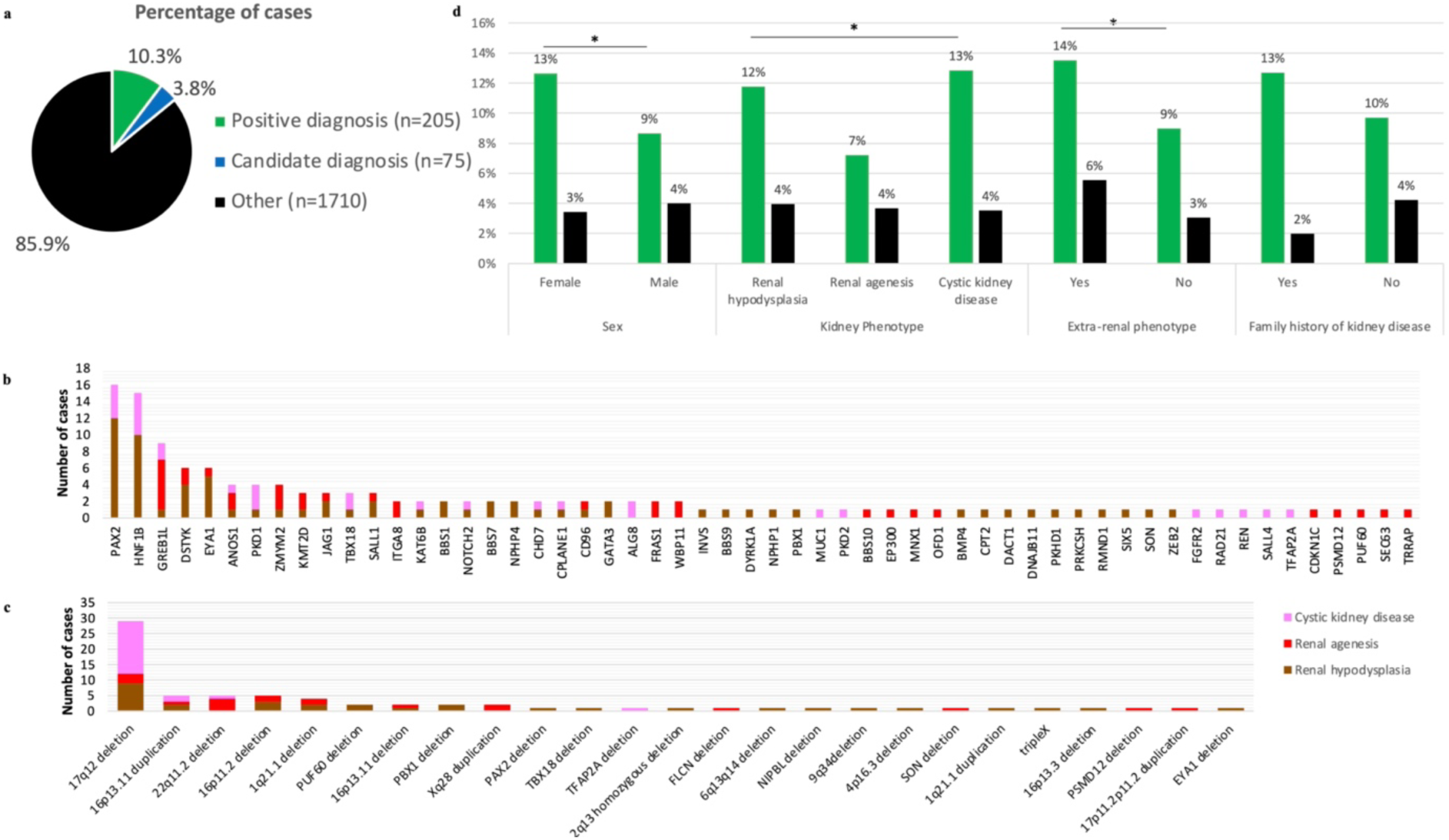
Diagnostic analysis in 1,990 individuals. (a) Percentages of cases with diagnostic findings (green) and a candidate diagnosis (blue: VUS-high in a known gene, P/LP in a candidate gene or candidate structural variant) and those without a diagnostic or candidate variant (black); (b) Number of cases with an LP/P variant diagnostic or candidate finding (cases with 2 variants were counted once, with the variant most likely associated with their form of KA) per gene and KA; (c) Number of cases with a diagnostic or candidate structural variant per genomic area and KA (pink: cystic kidney disease, red: renal agenesis and brown: renal hypoplasia). (d) Proportion of cases with diagnostic or candidate diagnostic findings based on their characteristics (sex, kidney phenotype, extra-renal phenotype, and family history). The statistical significance of the differences between groups only included individuals with diagnostic variants compared to those without candidate variants (n=1710). Asterisks (*) indicate statistical significance (p < 0.05) between groups.

**Table 1:** Characteristics of the cases included in the study.

|  |  | N | Proportion |
| --- | --- | --- | --- |
| Total |  | 1,990 |  |
| Sex | Female | 814 | 41% |
|  | Male | 1,176 | 59% |
| Participants' enrollment location or origin | Poland | 842 | 42% |
|  | Italy | 536 | 27% |
|  | Macedonia | 159 | 8% |
|  | Columbia University Irving Medical Center | 132 | 7% |
|  | Chronic Kidney Disease in Children Study (CKiD) | 112 | 6% |
|  | Deciphering Developmental Disorders (DDD) | 97 | 5% |
|  | Netherlands | 52 | 3% |
|  | Croatia | 37 | 2% |
|  | Other | 23 | 1% |
| Genetic ancestry (PCA based) | Europe | 1,736 | 87% |
|  | South America (Latino/Hispanic) | 70 | 4% |
|  | Africa | 54 | 3% |
|  | Asia | 17 | 1% |
|  | Admixed | 97 | 5% |
| Kidney phenotype | Hypoplastic or dysplastic kidney disease | 858 | 43% |
|  | Kidney agenesis | 735 | 37% |
|  | Multi-cystic dysplastic kidney disease | 397 | 20% |
| Additional lower urinary tract phenotypes | Including VUR, ureter, bladder and urethra anomalies | 371 | 19% |
| Extra-renal phenotypes (not mutually exclusive) | Congenital Heart Disease | 178 | 9% |
|  | Intellectual disability and developmental delay | 108 | 5% |
|  | Genital | 97 | 5% |
|  | Gastro-intestinal | 105 | 5% |
|  | Facial dysmorphology | 42 | 2% |
|  | Eye | 47 | 2% |
|  | Ear (including hearing impairment) | 45 | 2% |
|  | Endocrine | 65 | 3% |
|  | None reported <sup>a</sup> | 1413 | 71% |
| Study Cohort | Trios | 248 | 12% |
|  | Singletons (case-control analysis) | 1,742 | 88% |
|  | Singletons (single variants analysis) | 1,566 | 79% |

<sup>a</sup> Extra-renal presentations were not consistently collected in different cohorts.

Autosomal dominant disorders accounted for the majority of diagnoses (n=180, 87.8%), followed by autosomal recessive disorders (n=20, 9.8%) and X-linked disorders (n=5, 2.4%).

Amongst the 180 probands with dominant diagnoses, family information was available for 25; of those, 14 (56%) had de-novo variants, 6 (24%) inherited the variant from a parent with KA, and 5 (20%) inherited the variant from a parent without reported KA.

Overall, we observed a higher diagnostic rate in females (13% versus 9% in males, chi- square p-value= 4.6x10^-3^, **Fig. 1d**) and in those with extra-renal anomalies (14% versus 9%, chi- square p-value= 1.28x10^-3^). We also observed a higher diagnostic rate in probands with cystic dysplastic kidney diseases (13% versus 7% in probands with renal agenesis, chi-square p-value= 1.9x10^-3^). We only observed a trend for increased diagnostic rate based on reported family history of kidney disease (13% versus 10%, chi-square p-value= 0.11).

### Enrichment for de-novo variants in constrained genes in trios with Kidney Anomalies

A total of 370 *de-novo* variants were identified in 248 KA trios, and the number of *de-novo* variants per trio ranged from 0 to 12. The distribution of *de-novo* variants per trio was consistent with prior reports (**Fig. S1**). While there was no enrichment of de-novo synonymous variants, we observed a 1.85-fold enrichment of de-novo loss-of-function variants (LoF, FDR q-value=3.42 x10^-4^, **Table S4**), and a 1.36-fold enrichment of de-novo missenses (FDR q-value =1.54 x10^-5^) in cases compared to expectations. Of the 291 non-synonymous *de-novo* variants, only 8 were in genes known to be associated with dominant forms of KA (*HNF1β, KAT6B, PAX2, PSMD12, SON, NIPBL, PUF60, GREB1L*). The enrichment for *de-novo* variants was more pronounced for constrained genes (pLi>0.9, LOEUF<0.35, misZ>3.09), particularly those that are highly expressed during kidney development. When restricting the analysis to constrained genes highly expressed in human nephron-progenitor cells (NPCs), we observed a 7.64-fold enrichment for LoF variants (FDR q-value=8.52 x10^-5^), and a 3.78-fold enrichment for missense variants (FDR q-value=1.35 x10^-3^). Similarly, we observed a significant enrichment of LoF and missense variants in constrained genes highly expressed during early mouse kidney development (**Table S4**).

### Enrichment of rare variants in constrained genes by gene-based collapsing analysis

We also performed gene-based collapsing analysis in 1,742 unrelated probands with KA (not included in the trio analysis) and 22,258 genetically matched controls. Under the LoF model, two genes *HNF1β* and *PAX2*, both known to be associated with KA, reached exome-wide significance (p-value<10^-6^, **Fig. 2a**). No other genes reached exome-wide significance in any other models (**Fig. S2 and Fig. S3**). When we restricted the genome-wide analysis to constrained genes (pLi>0.9, LOEUF<0.35), we observed a significant enrichment of LoF variants (Odds Ratio OR_(LoF)_= 1.48; FDR q-value=4.03x10^-9^) and missense variants (OR= 1.26; FDR q-value= 4.29x10^-5^) (**Fig. 2b, Table S5**). The enrichment was slightly more pronounced when we restricted the analysis to constrained genes that are highly expressed in NPCs (OR_(LoF)_=1.70, FDR q-value= 1.12x10^-4^; OR_(Mis)_ = 1.32, FDR q-value= 2.7x10^-3^). We also noted that significantly more cases than controls harbored three or more LoF or missense qualifying variants in constrained genes (36% of cases vs 24% of controls, Pearson’s Chi-squared test p-value=1.7x10^-40^, **Fig. 2c, Fig. S4 and Table S6**). The cases with three or more qualifying variants in constrained genes were likelier to have extra-renal anomalies and complex urogenital phenotypes (11% of cases with extra-renal anomalies vs 8% of cases with unknown or no extra-renal anomalies, Pearson’s Chi-squared test p-value=1.3x10^-2^, **Fig. 2d**).

**Figure 2:**
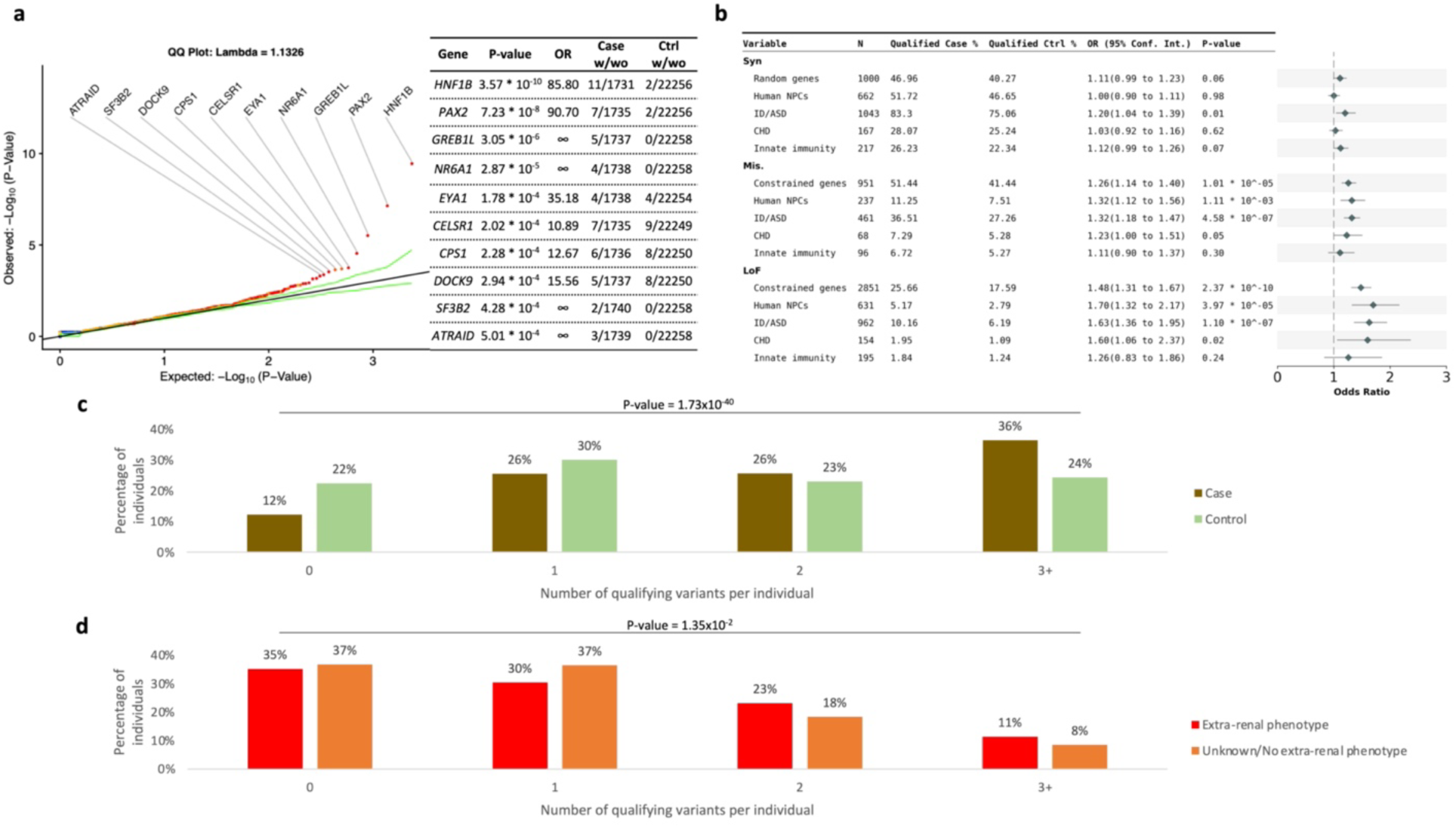
Gene-burden and gene-set enrichment analyses. (a) Quantile-Quantile probability plot of the p-values generated by the gene-burden analysis focused on qualifying LoF variants. List of the 10 most significant genes. (b) Forest plot depicting the gene-set analysis using six gene sets based on three gene burden analyses. (c) Comparison of the number of cases (brown) and controls (green) with 0,1,2 or 3+ qualifying variants (LoF and predicted deleterious missense variants) per individual in constrained genes. A significant difference was observed (p-value= 1.73x10^-40^). (d) A significant difference was observed when comparing cases with extra-renal phenotypes and cases without known extra-renal phenotypes with 0,1,2 or 3+ qualifying variants in constrained genes (p-value=1.35x10^-2^). CHD: congenital heart disorder, ID/ASD: intellectual delay and autism spectrum disorder, LoF: Loss-of-function variants, mis: missense variants, NPCs: Nephron Progenitor Cells, OR: Odds Ratio, syn: synonymous variants. Predicted deleterious missense variants: subRVI domain score percentile <50% and predicted damaging based on at least 2 of the following criteria: (1) REVEL>0.5; (2) PrimateAI>0.8; (3) polyphen possible/probable/unknown.

### Enrichment for rare damaging variants in genes associated with intellectual disability/autism spectrum disorder trio and case-control analyses

We searched for rare damaging variants in genes associated with ID/ASD and CHD. There was no significant enrichment in genes in the innate immune system, used as negative control. However, we detected an excess of de-novo LoF variants in constrained genes associated with ID and/or ASD both in the trios (4.11-fold enrichment, FDR q-value= 3.35x10^-4^, **Table S4**) and in the case-control dataset (LoF: OR_(LoF)_=1.63, FDR q-value = 9.18x10^-7^; OR_(Mis)_= 1.32, FDR q-value = 2.06x10^-6^, **Table S4**). Combining the trio and case-control analyses we confirmed the significant enrichment of variants in constrained genes, genes expressed in NPCs, genes associated with ID/ASD, and genes associated with CHD (**Table 2**).

**Table 2:** Meta-analysis of case-control and de-novo gene sets analyses.

| Gene-set | Nb of genes | Model | Case-control analysis p-value | De novo analysis p-value | Fisher p-value | FDR q-value |
| --- | --- | --- | --- | --- | --- | --- |
| Constrained Genes | 500 | Syn | 0.06 | 0.87 | 0.21 | 0.24 |
| | 2,850 | LoF | $2.37 \times 10^{-10}$ | $3.25 \times 10^{-5}$ | $2.58 \times 10^{-13}$ | $3.87 \times 10^{-12}$ |
| | 952 | Mis. | $1.01 \times 10^{-5}$ | $2.01 \times 10^{-3}$ | $3.80 \times 10^{-7}$ | $1.43 \times 10^{-6}$ |
| Human NPCs (18 weeks) | 663 | Syn | 0.98 | 0.58 | 0.89 | 0.9 |
| | 632 | LoF | $3.97 \times 10^{-5}$ | $1.42 \times 10^{-5}$ | $1.26 \times 10^{-8}$ | $6.30 \times 10^{-8}$ |
| | 238 | Mis. | $1.11 \times 10^{-3}$ | $2.26 \times 10^{-4}$ | $4.06 \times 10^{-6}$ | $1.01 \times 10^{-5}$ |
| Intellectual disability or Autism Spectrum Disease | 1,043 | Syn | 0.01 | 0.85 | 0.05 | 0.075 |
| | 962 | LoF | $1.10 \times 10^{-7}$ | $2.23 \times 10^{-4}$ | $6.24 \times 10^{-10}$ | $4.68 \times 10^{-9}$ |
| | 461 | Mis. | $4.58 \times 10^{-7}$ | 0.09 | $7.64 \times 10^{-7}$ | $2.29 \times 10^{-6}$ |
| Congenital Heart Disease | 286 | Syn | 0.6 | 1 | 0.9 | 0.9 |
|  | 267 | LoF | 0.02 | 0.03 | 0.01 | 0.019 |
| | 97 | Mis. | 0.05 | 0.02 | $7.30 \times 10^{-3}$ | 0.016 |
| Immune | 217 | Syn | 0.07 | 0.71 | 0.20 | 0.24 |
|  | 195 | LoF | 0.24 | 0.06 | 0.08 | 0.11 |
|  | 96 | Mis. | 0.30 | 0.02 | 0.04 | 0.067 |

LoF: Loss of Function variants (stop-gained, stop-lost, start-lost, splice-site and frameshift variants), Mis.: missense variants, Syn: synonymous variants.
Constrained genes only (Constrained: <sup>a</sup> for LoF: $pLi > 0.9$ and $oe\_lof\_up\_ < 0.35$ , <sup>b</sup> for missenses: $misZ > 3.09$ )
NPCs: Top decile expression in Nephron Progenitor Cells at 18 weeks of gestation, constrained genes only
ASD: Genes associated with Autism Spectrum Disorder extracted from the AutismDB, constrained genes only
CHD: genes associated with Congenital Heart Disease, constrained genes only

These findings were further supported by an analysis of clinical phenotypes. Of the 177 probands with diagnostic variants, 60 had variants in genes associated with KA and developmental delay, and despite the paucity of clinical information, 11 (18%) of those had reported intellectual disability (ID) or developmental delay. Similarly, 50 probands had variants in genes associated with KA and CHD, and 8 (16%) of them had reported CHD (**Table S3**).

### Two novel genes, *NR6A1* and *ARID3A*

The trio and collapsing analyses identified 40 novel candidate genes for KA prioritized based on predicted deleteriousness, mutational constraint, known association with ID/ASD or CHD, enrichment for collapsing analysis (OR>10) or de-novo status (**Table S7**). In the genes *SSBP2, XPO1, MAP4K4, NR6A1,* and *ARID3A*, at least two cases were identified with loF variants while no LoF variants were identified in controls (**Table S7**). *NR6A1* (nuclear receptor subfamily 6 group A member 1) and *ARID3A* (AT-rich interaction domain 3A) with respectively four and three LoF variants in cases, were both confirmed as new disease genes by identification of additional independent probands with KA and related phenotypes.

### NR6A1

Review of sequencing data from Columbia CAKUT cases not included in the discovery cohort identified 6 additional cases with *NR6A1* predicted deleterious variants (predicted splice variants in two cases and predicted deleterious missense variants with REVEL>0.8 in four cases, **Table 3**, **Fig. 3**). One of these cases harbors an R92W variant that segregates in additional affected relatives (**Fig. 3b**). Structural analysis predicts R92W is highly deleterious because it would abrogate the formation of high affinity salt bridges with the DNA phosphate backbone and would cause clashes within the DNA binding domain (**Fig. 3c**, **Fig. 3d**). The Columbia KA proband with the R92W variant also has an eye coloboma but no other variants in genes causing eye defects (**Fig. 3b**). Through collaboration with the NIH, we identified two unrelated individuals ascertained for developmental eye phenotypes who also had KA and harbored pathogenic *NR6A1* variants (a large deletion and the R92W variant, see related manuscript by Neelathi et al.). Thus, the R92W variant was identified in three unrelated probands with KA and eye abnormalities in the Columbia and NIH cohorts. We also identified an additional *NR6A1* LoF variant through collaboration with UMC Utrecht. The UMC Utrecht case had CAKUT In summary, we identified *NR6A1* predicted deleterious variants in 10 cases in the Columbia cohort (including 6 LOF and 1 *de-novo;* 8 with KA and 2 with KA and eye anomalies), and 3 additional cases in external cohorts (including 2 LoF variants; 2 with KA and eye anomalies and 1 with eye anomalies), identifying *NR6A1* as a novel gene associated with kidney, eye, and other congenital anomalies.

**Figure 3:**
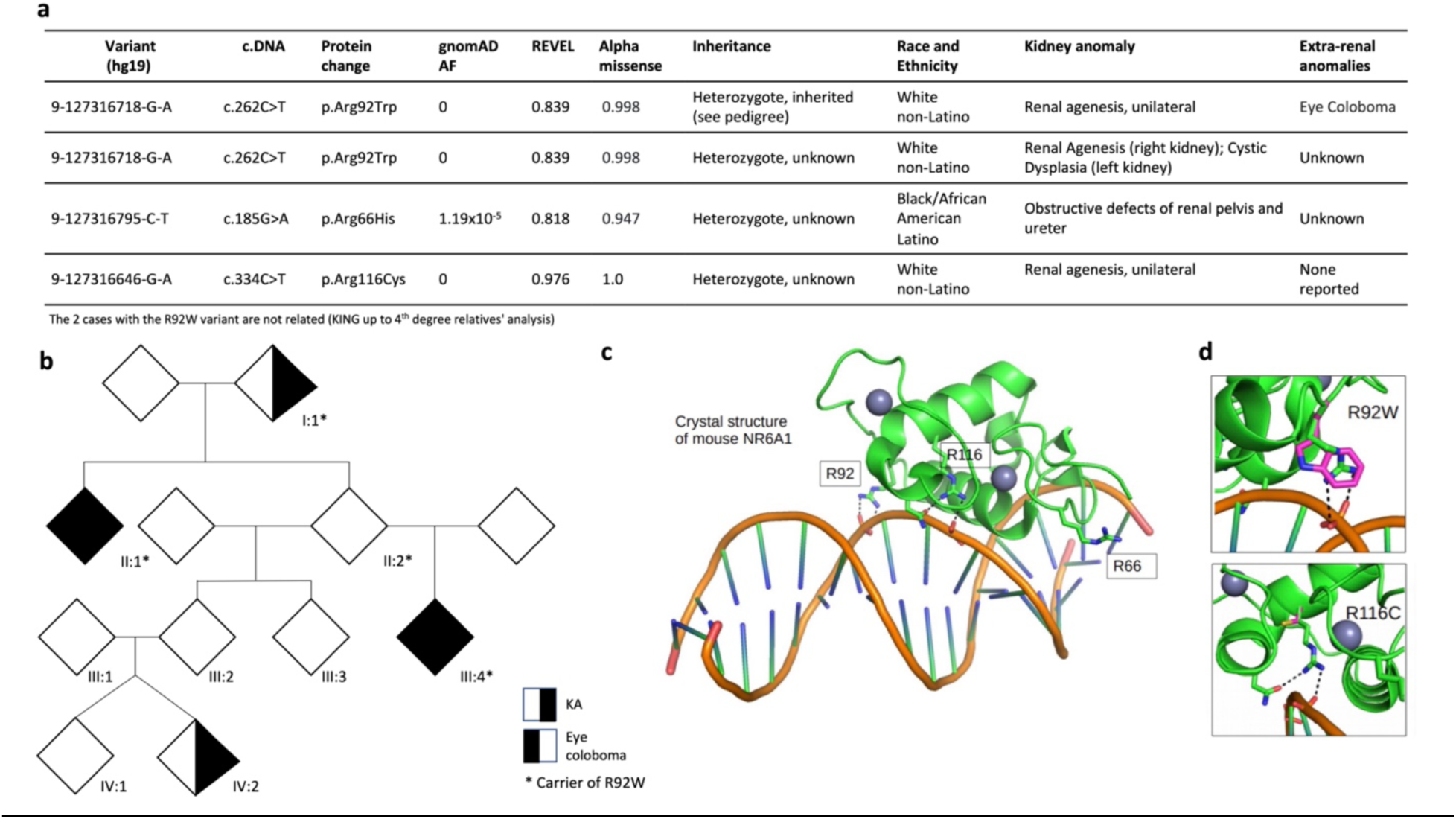
Missense variants in *NR6A1*. (a) Four independent cases with predicted deleterious missense variants in *NR6A1*. (b) Family carrying the p.Arg92Trp (R92W) variant in *NR6A1.* (I:1 Pelvic and small kidney, II:1 Renal agenesis and Eye coloboma, III:3 Renal agenesis and Eye coloboma, IV:2 Renal agenesis and unknown genetic status). (c) Crystal structure of mouse NR6A1 and location of the three amino acids in which missense variants were identified in the four cases. (d) Modeling of the potential impact of two of the three missense variants on the protein structure (R92W and R116C are modeled on the NR6A1 crystal structure PDB ID: 5KRB) The potential structural consequences of R66H could not be determined from the published NR6A1 structure.

**Table 3:**
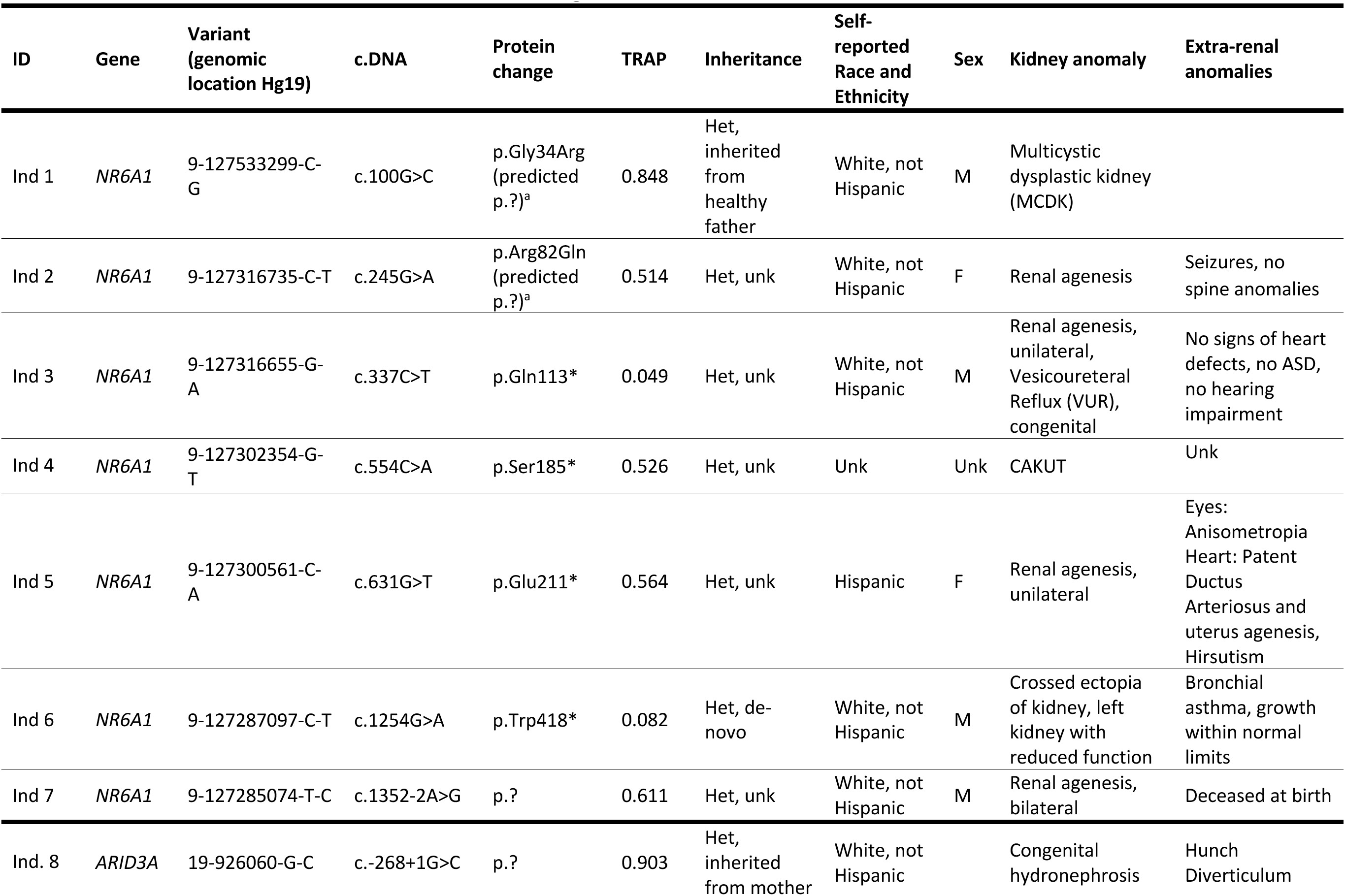
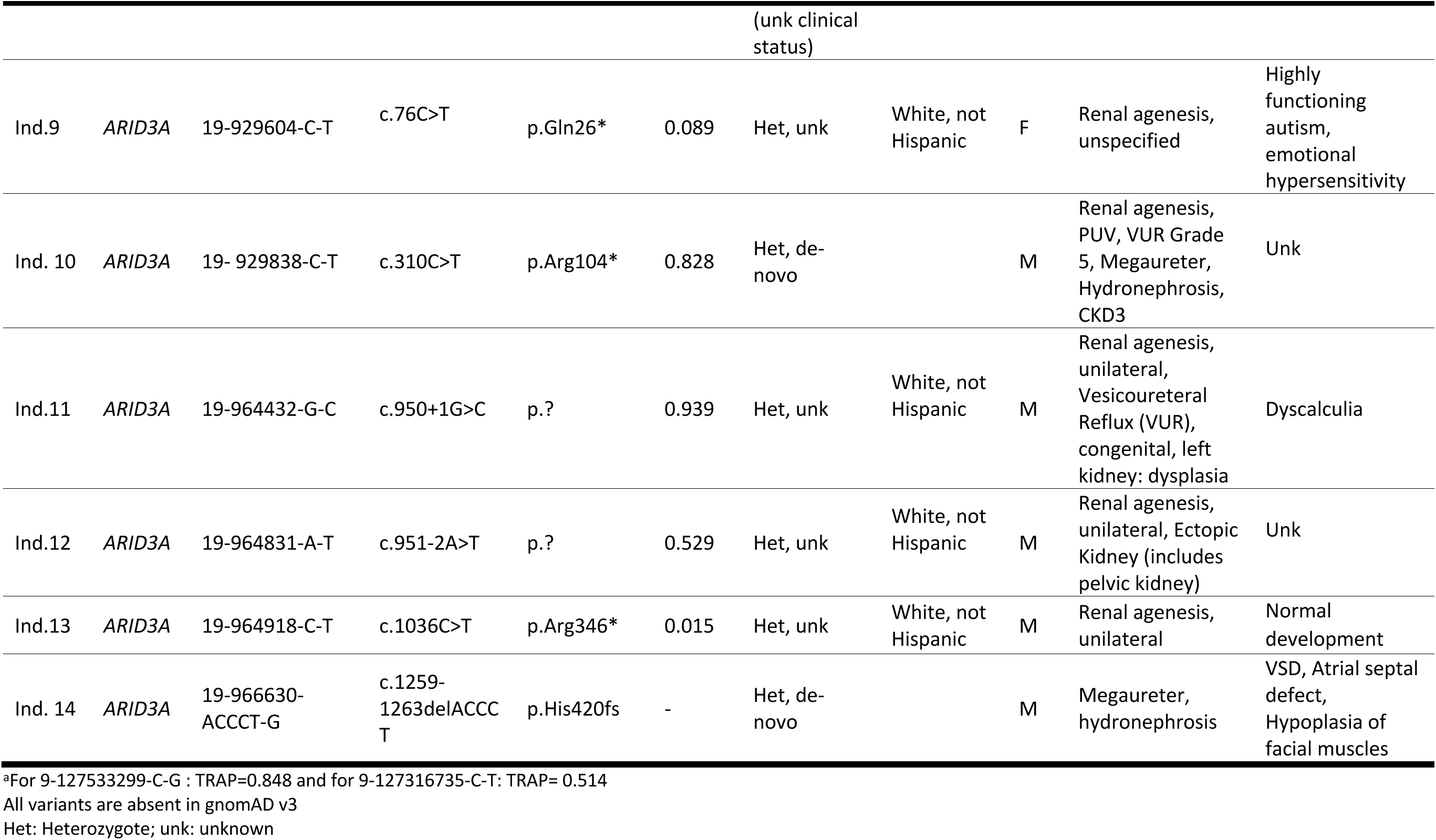
Predicted Loss-of-function variants in two novel genes: *ARID3A* and *NR6A1*.

### ARID3A

In addition to the three LoF variants in our discovery cohort (**Table 3**), a review of our biobank identified two additional KA cases with *ARID3A* LoF variants. Through collaboration with Boston Children’s Hospital, we identified two additional individuals with LoF in *ARID3A,* both confirmed as *de-novo* variants. One individual displays VUR Grade 5, Solitary Kidney, PUV, Megaureter, Hydronephrosis, and chronic kidney disease stage 3. The other individual displays Megaureter, hydronephrosis, VSD, Atrial septal defect, and Hypoplasia of facial muscles. Altogether, we identified 7 predicted LoF in *ARID3A* in patients with kidney and genitourinary defects. The finding of seven independent LoF variants, including two DNV, identifies *ARID3A* as a novel haploinsufficient CAKUT gene. A phenotypic review indicated that patients with *ARID3A* LoF variants have multiple genitourinary defects, and a few had extra- renal abnormalities. Pathway analysis demonstrated that *ARID3A1* and *NR6A1* are co-expressed with *SALL4*, a gene known to cause KA (**Fig. S5**).

### Low-frequency risk variants

We also performed an exome-wide single variant association study for low-frequency risk variants in genes associated with known to be associated with KA (**Table S2**). We identified four suggestive associations in genes known to cause autosomal dominant or recessive KA (*DSTYK, KMT2D, PKHD1,* and *SDCCAG8)*, including the low-frequency *DSTYK* variant with a prior association with CAKUT (**Table S8**).^23^

### Discussion

In this cohort of 1990 individuals with KA, the diagnostic, trio, and case-control analyses point to a wide number of genes for KA. We identified diagnostic variants in 57 genes and 25 structural variants. Combined with the candidate diagnostic variants (54 genes and four structural variants), the overall diagnostic rate of 14.1% in this cohort falls in the previously reported range for congenital disorders^6^. *HNF1β* and *PAX2* accounted for a larger fraction of diagnoses and therefore reached genome-wide significance in the case-control analysis, while other genes known to be associated with KA each contributed a small fraction of cases, highlighting the long tail in the distribution of rare genetic diseases that cause KA. The trio and case-control analyses also demonstrated a significant excess of ultrarare deleterious variants, mostly in constrained genes expressed during early kidney development that are not known to be associated with KA, suggesting many yet-to-be-identified genetic causes that would require larger cohorts for discovery. Moreover, KA patients harbored a significant burden of deleterious variants in ID/ASD genes, and the candidate diagnoses also pointed to KA as an expansion phenotype for many known CHD and ID/ASD syndromes. These data are consistent with our prior studies showing significant overlap between CNV disorders underlying KA, CHD, and ID/ASD and that KA patients with pathogenic CNVs often have undetected or subclinical neurodevelopmental phenotypes. In this cohort, we similarly found that the KA patients frequently have other cardiac or neurodevelopmental comorbidities that are clarified by the genetic analysis. Altogether, these findings indicate shared architecture between KA, CHD, and ID/ASD and point to the potential for phenotypic expansion for many developmental disorders. Hence, a cross-phenotype analysis of developmental disorders may identify new genetic disorders and potential modifiers that determine the spectrum and severity of defects.

The reasons for variable severity of clinical phenotypes in KA patients are unknown; genetic, environmental, or developmental factors have been invoked.^24^ We found that 5% KA cases harbored two or more ultrarare deleterious variants in constrained genes, and these individuals had a higher likelihood of presenting with multiple developmental phenotypes. This finding is supported by our prior study where we similarly found that KA patients with extrarenal phenotypes often harbor multiple rare, gene-disrupting CNVs.^6^ In addition to a higher burden of ultrarare variants, we identified several low-frequency risk variants in *DSTYK, KMT2D, PKHD1,* and *SDCCAG8*, which may modify the severity of disease, consistent with prior studies demonstrating that common variants also influence the risk of developmental phenotypes. Taken together, these data suggest oligogenic or polygenic determination for some KA cases, and variation in genetic background as a potential contributor to phenotypic severity.

We identified two novel genes for KA, *NR6A1* and *ARID3A*. We found multiple probands and families with *NR6A1* mutations presenting with kidney and eye malformations and a few other developmental phenotypes. ***NR6A1*** encodes the protein germ cell nuclear factor (GCNF), also known as RTR (Retinoid Receptor-Related Testis-Associated Receptor). *Nr6a1* is expressed at 15 days post coitum (dpc) in mice in the ureteric tip and the cap mesenchyme (TS19-TS28), and in the early tubule and renal interstitium^25^. *Nr6a1*^−/−^ embryos have abnormal optic vesicle morphology, neural tube closure, and additional developmental anomalies leading to their demise at 10.5 dpc. Because kidneys develop after 12 dpc, conditional inactivation in cells important for kidney development will be required to study the function of *Nr6a1* on kidney development. The role of *NR6A1* variants in causing eye and kidney defects was confirmed by the identification of independent cases ascertained for eye anomalies (Neelathi and colleagues). The incomplete penetrance and expressivity identified in carriers of *NR6A1* variants is a common phenomenon in autosomal dominant diseases^26^.

We identified a total of 5 individuals with unilateral renal agenesis and two individuals with hydronephrosis with rare predicted deleterious LoF variants in ***ARID3A***, and *t*wo of those variants occurred *de-novo*. Interestingly, a review of 13 cases with a 19p13.3 microdeletion that encompasses *ARID3A* reported 3 cases (23%) with unilateral renal agenesis.^27^ *ARID3A* encodes a DNA-binding protein highly expressed in E14.5 murine kidneys^25^. Inactivation of *Arid3a* leads to embryonic lethality at E12.5 for most mice due to impaired hematopoiesis, but the few surviving *Arid3a* null mice display abnormal cell proliferative and structural abnormalities in their kidneys.^28,29^ In *Xenopus*, ARID3A binds to regeneration signal-response enhancers, reduces the levels of the constitutive heterochromatin marker histone three lysine nine trimethylation, promotes cell cycle progression, and causes the outgrowth of nephric tubules.^30^ In addition, an *Arid3a*^−/−^ kidney mouse cell line, KKPS5, was reported to spontaneously develop into multicellular nephron-like structures in vitro, which can form mouse nephron structures if engrafted into immunocompromised medaka mesonephros.^31^ Interestingly, we found that *ARID3A* is co-expressed with *NR6A1* and *SALL4* and is predicted to function in a common transcriptional module.^32–34^ Taking together, these data suggest that *ARID3A* plays a regulatory role in nephrogenesis, potentially by balancing cellular proliferation. Hence, other co-expressed genes in the *ARID3A* transcriptional module may be candidates for kidney developmental disorders.

Our study has multiple clinical and research implications that chart a path forward for genetic studies of KA. Our data highlight the significant genetic heterogeneity of KA and expand the list of potential diagnostic genes for this disorder. The genetic overlap with other developmental disorders suggests that a diagnostic workup of KA patients should include a broader gene list, and KA should be considered as an expansion phenotype for genetic disorders associated with cardiac, eye or brain developmental disorders. In addition, KA patients should be screened for extrarenal developmental phenotypes that may have been missed during initial evaluation, particularly neurodevelopmental phenotypes which may become evident in later years. Conversely, KA genes should be considered in the workup for other developmental phenotypes. The determinants of variable clinical expression of kidney malformations suggest a role for genetic modifiers or redundant developmental programs that influence the ultimate clinical outcome. While the diagnostic and gene burden analysis nominated several candidate genes such as *SPBP2*, *XPO1,* or *MAPK4*, the excess number of rare variants in conserved genes detected in the trio and case-control analyses also point to the existence of many additional KA genes that will require larger cohorts for discovery. Finally, the genes for kidney malformations may also point to environmental factors that can disrupt developmental programs. For example, several genes implicated in KA such as *RET* or *GREB1L* interact with retinoic acid signaling, which is important for organogenesis.^35,36^ Similarly, disruption of the gut microbiota in obese mice may influence *ARID3A* binding to its targets by altering specific metabolites.^37^

Study limitations include incomplete clinical information about the spectrum of developmental phenotypes. In addition, although this is the largest genetic study of KA patients to date, a still larger sample size will be needed to comprehensively assess the spectrum of genes and variants contributing to KA. Detailed phenotyping, consistent collection of family history, and larger cohorts would allow a significant increase in both diagnostic yield and identification of novel disease-causing genes that may have smaller effect sizes or act under a recessive model. Collaborative studies combining patients with KA, CHD, ID, and ASD will also help overcome the challenge associated with genetic heterogeneity and limited penetrance and also enable long-term assessment of functional status, clinical complications, and reproductive outcomes.

### Materials and Methods Subjects

The diagnosis of KA was made by nephrologists or urologists based on pertinent imaging data such as renal ultrasound, MRI, or CT scans. The study cohort was composed of 1781 individuals and 176 parents recruited for genetic studies of kidney disease at Columbia University and collaborating institutions from Poland, Italy, Macedonia, the Netherlands, Croatia, and other countries (**Table 1**). In addition, the study includes 112 KA probands from the Chronic Kidney Disease in Children (CKiD) study^38^ and 277 samples from the Deciphering Developmental Delay (DDD) study^16^, including 90 trios of probands with KA and parents without KA and seven probands whose parents have KA. Probands included 858 cases with hypoplastic or dysplastic kidney disease, 735 cases with kidney agenesis, and 397 cases with multicystic dysplastic kidney disease (Table 1). The majority of cases were of European ancestry (87%) and were not reported to have extra-renal phenotypes (71%).

Of the 1,990 probands, 248 were analyzed as trios (no family history, **Fig. S6)**. Family relationships were confirmed using KING software.^39^ The sequence data from 1,742 unrelated (KING V1.4: up to third-degree, kingship coefficient > 0.0884) probands with KA who were not included in the trio analysis were compared to data from 22,258 unrelated controls matched based on genetic ancestry (**Table S1**). The controls were enrolled in studies unrelated to KA or as controls and were available through the Columbia Institute for Genomics Medicine (IGM), all consented to have anonymized sequence data available for secondary genetic analysis. These included 2,814 individuals with immune-mediated forms of kidney diseases (biopsy documented IgA nephropathy or C3 glomerulopathy), 10,114 individuals with disorders unrelated to kidney disease (e.g. amyotrophic lateral sclerosis), and 9,330 individuals enrolled as controls or healthy family members to diverse studies. Approval for human research was obtained from the Institutional Review Board of Columbia University Medical Center, as well as other relevant Ethics Review Boards at collaborating institutions.

### Exome Sequencing

Exome sequencing was performed on 2,290 samples (1,893 probands and 316 parents) through the Yale Center for Mendelian Genomics, the New York Genome Center, and the IGM, using different exome kits (**Table S9)** at a minimum average depth of 30X to a maximum of 250X as described.^21,40^ All samples had a minimum of 90% coverage at 10X across the Consensus Coding Sequence (CCDS) regions. In addition, the CRAM files from the DDD study (Agilent V5 exome kit) were downloaded and converted into FASTQ files.^16^ All probands, parents, and controls’ sequence data were processed using the same bioinformatic pipeline. FASTQ data were processed and aligned to hg19/GRCh37 using DRAGEN and GATK haplotype caller, and variants were called using the Genome Analysis Toolkit (GATK) v3.6 best practices. Analyses were performed using the in-house Analysis Tool for Annotated Variants for retrieving, annotating, and filtering variants in large cohorts.^41^

### Diagnostic analysis

We used a combination of automated and manual curation to generate a gene list for diagnostic analysis. Human Phenotype Ontology (HPO) terms associated with KA were utilized to capture a comprehensive list of relevant genes.^42^ Genes associated with a single HPO term underwent a manual curation of the literature to determine gene-phenotype associations. The list was also annotated for the presence of extra-renal manifestations.

The single-nucleotide variants and small insertions and deletions were classified according to the American College of Medical Genetics and Genomics (ACMG) guidelines for clinical sequence interpretation.^43^ Structural variants were identified with either or both microarray data and exome data. To identify copy-number variants with microarray data, samples were genotyped on various Illumina arrays (**Table S9**) and analyzed as previously described.^6,44^ To identify structural variants in exome data we used XHMM.^45^ As XHMM does not readily differentiate between homozygous and heterozygous deletions, we only analyzed deletions in genes known to be associated with dominant forms of KA.

When available, parental inheritance data was used to determine the variants’ phase and de-novo status. Variants were classified as pathogenic (P), likely pathogenic (LP), or variants of uncertain significance highly suspicious to become pathogenic because of phenotype association (VUS-high).^22^ Only variants that matched the inheritance pattern of the associated condition were considered. P/LP variants and full gene deletions in genes known to cause KA were considered diagnostic. In the probable genes, only P/LP variants that matched the inheritance pattern of the associated condition are reported.

### *De-novo* variants’ identification

Variants were annotated using Ensembl canonical transcripts. In cases with multiple transcripts, the most deleterious effect was included. *De-novo* variants were defined as variants present in the proband (Read Depth≥10, Alternate allele depth≥ 3, QUAL_snv_ ≥ 50, QUAL_indel_ ≥ 300, GQ ≥ 20, proportion of missing alleles less than 20%, Alternate read percentage 20-80%). The stringency of these quality filters was tested using inherited variants in the same trios, demonstrating that they removed less than 1% of inherited variants. Finally, only rare variants based on the Genome Aggregation Database^46^ (gnomAD v2.1.1 global allele frequency (AF) ≤ 10^-5^ and IGM AF≤ 10^-4^) absent in the parents of the trios analyzed were retained. All variants were then manually curated using the Integrative Genomics Viewer (IGV) interactive software tool.

### De-novo enrichment analysis

De-novo analysis was restricted to functional *de-novo* variants: LoF variants (LoF: canonical splice-site, stop-gain, stop-loss, start-lost, frameshift insertions & deletions), missense variants, and protein-altering (LoF and Missense) variants. To estimate the probability of a DNV with each one of those effects, we used denovolyzeR^47^ and the updated mutation table from denovoWEST.^48^ The p-value generated by denovolyzeR was obtained from a Poisson test. For the genome-wide analysis, a Bonferroni corrected p-value threshold at *α*=0.05 is used, i.e. 1.3x10^-6 47^.

### Predefined gene-sets

Our primary analysis focused on comparing the burden of *de-novo* variants or ultra-rare variants in two sets of mutation intolerant genes as predicted by gnomAD: one set of genes with a high probability of being loss-of-function (LoF) intolerant (pLI score > 0.9, LOEUF>0.35 ; n=2,851 genes, Bonferroni corrected p-value=8.8x10^-6^), and one set of genes intolerant to missense variation (missense Z score > 3.09 , n=952 genes, Bonferroni corrected p-value=2.6x10^-5^). We also analyzed a subset of those two gene-sets: genes highly expressed (top decile) in nephron-progenitor cells (NPCs) in 18 weeks human embryonic kidneys^49^ (n=610 genes for the LoF intolerant set and n=228 genes for the missense-intolerant set) and genes highly expressed (top decile) during early murine kidney development at E15.5 (n=434 genes for the LoF intolerant set, Bonferroni corrected p-value=5.8x10^-5^and n=189 genes for the missense-intolerant set, Bonferroni corrected p-value=1.3x10^-4^). The genes highly expressed during early murine kidney development were identified using bulk RNAseq assays. Embryos were harvested at 15.5-days post-conception (dpc). Kidneys were then microdissected out of the embryos and stored in RNAlater (Invitrogen, ThermoFisher Cat no. AM7020) for future pooling and subsequent total RNA extraction using TRIzol (ThermoFisher Cat no. 15596026).

Specifically, each biological replicate (“sample”) that was sequenced represents RNA from a pool of kidneys (2 embryos, or 4 kidneys total, per sample; final n = 3). Samples with a RIN (RNA Integrity Number) score of 9.0 or greater were then submitted to the Columbia Genome Center Sequencing Core for library generation and subsequent sequencing. Libraries were prepared subsequent to ribosomal RNA reduction (RiboZero) for mRNA purification. Samples were then sequenced on an Illumina NovaSeq 6000 (Illumina v4 chemistry) at a depth of 40 million paired-end reads.

We also created a set of genes associated with ID^50^ or ASD^51^ and subdivided the set into 962 genes intolerant to LoF variants (Bonferroni corrected p-value=2.6x10^-5^) and 461 genes intolerant to missense variants (Bonferroni corrected p-value=5.4x10^-5^). In addition, we analyzed a gene-set comprising 525 genes associated with congenital heart disease (CHD), compiled using existing clinical targeted panels offered to patients with CHD. The list was subdivided into a set of 154 genes intolerant to LoF variants (Bonferroni corrected p-value=6.5x10^-5^) and a set of 68 genes intolerant to missense variants (Bonferroni corrected p-value=1.5x10^-4^). As a negative control, we analyzed a Reactome gene-set comprising 1,124 genes associated with the innate immune system^52^ and subdivided the set into 195 genes intolerant to LoF variants (Bonferroni corrected p-value=1.3x10^-4^) and 96 genes intolerant to missense variants (Bonferroni corrected p-value=2.6x10^-4^). To adjust for the multiple testing, we calculated the FDR using the FDR function in the fuzzySim R package.^53^ The list of genes in each gene set can be found online (https://github.com/ColumbiaCPMG/genelists ).

### Case-control analysis

We performed a gene-burden genome-wide search for enrichment of “qualifying variants” in protein-coding genes in 1,742 KA compared to 22,258 controls.^54^ We restricted the analyses to variants within CCDS regions or the 2 bp canonical splice sites. Furthermore, we only considered variants that fulfilled all of the following QC criteria: >= 10x coverage of the site, quality score (QUAL) >= 50, genotype quality score (GQ) >= 20, quality by depth score (QD) >= 5, mapping quality score (MQ) >= 40, read position rank sum score (RPRS) >= -3, mapping quality rank sum score (MQRS) >= -10, Fisher’s strand bias score (FS) <= 60 (SNVs) or <= 200 (indels), strand odds ratio (SOR) <= 3 (SNVs) or <= 10 (indels), GATK Variant Quality Score Recalibration filter “PASS”, “VQSRTrancheSNP90.00to99.00”, or “VQSRTrancheSNP99.00to99.90”, alternate allele fraction for heterozygous calls >= 0.3. To control the differences in coverage, only variants covered in at least 70% of the case-control cohort and with a maximal 7% difference in coverage between cases and controls were included. To control for population stratification, we first performed Principal Component Analysis (PCA) on a set of predefined variants to capture population structure^55^. As described in Povysil et al. 2020, we used the Louvain method of community detection on the first 6 principal components (PCs) to identify 10 clusters divided based on genetic ancestry^54^. To assess the quality of the clusters we performed further dimensionality reduction using the Uniform Manifold Approximation and Projection (UMAP) and we performed the burden test for each gene in each cluster (**Table S10**). We then generated a combined P-value and odds ratio using the Cochran–Mantel–Haenszel test.^56^ To qualify, the variants’ allele frequency (AF) had to be less than 1×10^-5^ in the Genome Aggregation Database (gnomAD) popmax allele frequency and less than 1×10^-4^ in the IGM biobank. This AF was chosen as all diagnostic variants identified had a gnomAD MAF<1 × 10^-5^. Only LoF variants with high confidence based on LOFTEE^46^ and identified in regions with low regional allele frequency^57^ were “qualified”. Missense variants located in constrained domains based on a subRVI^58^ domain score percentile <50% and predicted damaging based on at least 2 of the following criteria: (1) REVEL^59^>0.5; (2) PrimateAI^60^ >0.8; (3) polyphen^61^ possible/probable/unknown were “qualified”. Two analyses were performed, one for LoF only and one for missenses only. After Bonferroni correction, genome-wide significance was identified as p-value<1.3x10^-6^. To confirm that this correction was adequate, we performed a case-control analysis of synonymous variants only and identified the lowest p-value=5.8x10^-5^. The number of qualifying LoF variants per sample in constrained genes was calculated to compare the distribution in cases and controls.

### Novel genes analysis

To determine the potential impact of rare variants on splicing, we added to the tools described above for the case-control analysis the TRAP score. ^62^

### Low-frequency variants analysis

To identify novel genetic risk factors for KA, we performed a single variant analysis of low-frequency variants (defined as gnomAD allele frequency ≤1%). To reduce the risk of genetic stratification, we only included unrelated cases (n=1566) and matched controls (n=13,031) with European genetic ancestry. We tested a total of 1,497 low-frequency variants in genes known to be associated with KA, identified in at least 10 individuals and covered in at least 90% of the cases and controls, gnomAD filter “PASS” and maximum 0.01 difference with gnomAD AF. Variants passed the same criteria as described in the case control analysis and the maximum allowed difference in coverage between cases and controls was 5%. Variant-level p-values were generated using a Fisher’s exact two-side test. As we ran a single model, after Bonferroni correction, we identified p-value ≤ 3.3x10^-5^ for genome-wide significant associations and a cutoff of OR>1.5 and p-value≤10^-3^ for suggestive associations. To determine the potential deleteriousness of low-frequency variants, we added to the tools described above for the case-control analysis the AlphaMissense score.^63^

### Pathway analysis

To assess whether the novel genes identified in this study co-express with any of the known KA genes, we used COXPRESdb.^64^ We first used the “CoExSearch” option to identify the top 300 genes co-expressed with each novel candidate gene. We then extracted from those 300 genes the known KA genes. We combined the lists obtained for each candidate gene and used the “NetworkDrawer” option using the automatic platform, Cytoscape, “draw PPIs” and either the “add a few genes” or “add many genes”.

## Data Availability

All data produced in the present study are available upon reasonable request to the authors

https://github.com/ColumbiaCPMG/genelists

https://ega-archive.org/studies/EGAS00001000775

## Acknowledgment

Donald E. Wesson Research Fellowship from the ASN Foundation for Kidney Research (HMR)

NDDK grants: Genetics of Human Renal HypoDysplasia (5R01DK080099-13, AGG), George M O’Brien Center (5U54DK104309-10, AGG), Advancing equitable implementation of genomic medicine in nephrology (5K01DK132495-02, HMR)

*The DDD study presents independent research commissioned by the Health Innovation Challenge Fund [grant number HICF-1009-003]. This study makes use of DECIPHER (*http://www.deciphergenomics.org*), which is funded by Wellcome [grant number WT223718/Z/21/Z]. See Nature PMID: 25533962 or* www.ddduk.org/access.html *for full acknowledgment. Source:* https://ega-archive.org/studies/EGAS00001000775

## The authors thank the following individuals or groups for contributing control samples

A Signature Program of CURE, Anna Mae Diehl, B. Grinton, C. Hulette, Croasdaile Village, D. Lancet, E. Davis, E. Nading, Farfel, G. Cavalleri, Heidi White, I. Scheffer, J. Burke, J. McEvoy, J. Samuels, K. Whisenhunt, Kenneth Schmader, Mamata Yanamadala, N. Delanty , S. Sisodiya, Shelley McDonald, A. Need, A. Poduri, C. Chen, C. Depondt, Carol Woods, Chia-siang Chen, Cynthia Moylan, D. Levy, Duke University Health System Nonalcoholic Fatty Liver Disease Research Database and Specimen Repository, E. Pras, Epilepsy Genetics Initiative, G. Nestadt, K. Welsh-Bomer, M. Gennarelli, M. Hauser, M. Sum, M. Walker, Maher, UCB funding, Manal Abdelmalek, N. Katsanis, "National Institute of Allergy and Infectious Diseases Center for HIV/AIDS Vaccine Immunology (CHAVI) (U19-AI067854), National Institute of Allergy and Infectious Diseases Center for HIV/AIDS Vaccine Immunology and Immunogen Discovery (UM1- AI100645), ", National Institute on Aging (R01AG037212, P01AG007232). ALSO include: Data collection and sharing for the WHICAP project (used as controls in this analysis) was supported by the Washington Heights-Inwood Columbia Aging Project (WHICAP, PO1AG07232, R01AG037212, RF1AG054023) funded by the National Institute on Aging (NIA) and by the National Center for Advancing Translational Sciences, National Institutes of Health, through Grant Number UL1TR001873. This manuscript has been reviewed by WHICAP investigators for scientific content and consistency of data interpretation with previous WHICAP Study publications. We acknowledge the WHICAP study participants and the WHICAP research and support staff for their contributions to this study, R. Buckley R. Wapner, S. Berkovic, S. Delaney, S. Palmer, S. Schuman, T.Young, the ALS Sequencing Consortium; the Washington University Neuromuscular Genetics Project, the Epi4K Consortium and Epilepsy Phenome/Genome Project, The Murdock study community registry and biorepository Pro00011196; Kristen Newby, V. Shashi, V. Shashi, Undiagnosed Diseases Network, Y. Wang

## Supplementary figures legends

**<u>Figure S1</u>: Distribution of trios by the number of de-novo variants identified in each of them.**

**<u>Figure S2</u>**: **Gene-burden analysis focused on predicted deleterious missense variants**. (a) Quantile-Quantile probability plot of the p-values generated. (b) List of the 10 most significant genes. Predicted deleterious missense variants: subRVI domain score percentile <50% and predicted damaging based on at least 2 of the following criteria: (1) REVEL>0.5; (2) PrimateAI>0.8; (3) polyphen possible/probable/unknown.

**<u>Figure S3</u>: Gene-burden analysis focused on LoF and predicted deleterious missense variants.**

(a) Quantile-Quantile probability plot of the p-values generated. (b) List of the 10 most significant genes. Predicted deleterious missense variants: subRVI domain score percentile <50% and predicted damaging based on at least 2 of the following criteria: (1) REVEL>0.5; (2) PrimateAI>0.8; (3) polyphen possible/probable/unknown.

**<u>Figure S4</u>: Comparison of the number of cases and controls by the number of rare variants per individual in constrained genes.** (a) LoF variants. (Pearson’s Chi-squared test, p-value= 8.91x10^- 19^) (b) Predicted deleterious missense variants (Pearson’s Chi-squared test, p-value= 1.15x10^-20^). LoF: Loss-of-function. Predicted deleterious missense variants: subRVI domain score percentile <50% and predicted damaging based on at least 2 of the following criteria: (1) REVEL>0.5; (2) PrimateAI>0.8; (3) polyphen possible/probable/unknown.

**<u>Figure S5</u>: Pathway analysis of genes co-expressed with *ARID3A* and *NR6A1***. (a) Table of the top genes co-expressed with *ARID3A* and/or *NR6A1* and associated with KA (Table S3). (b) Gene network using the genes in (a) and allowing NetworkDrawer to add many genes. (c) Gene network using the genes in (a) and only allowing NetworkDrawer to add a few genes.

**<u>Figure S6</u>: Study design**. The study includes sub-cohorts: one used for diagnostic analysis (orange), one used for de-novo analysis in trios (blue) and one used for case-control analysis (purple). The probands in the case-control analysis and the probands in the de-novo analysis do not overlap, and together they consist of all the probands included in the diagnostic analysis.

