## Supplemental figures S1-S6 and Supplemental tables S1, S4-S6, S8-S10 for "Exome-wide analysis of congenital kidney anomalies reveals new genes and shared architecture with developmental disorders"

### Figure S1

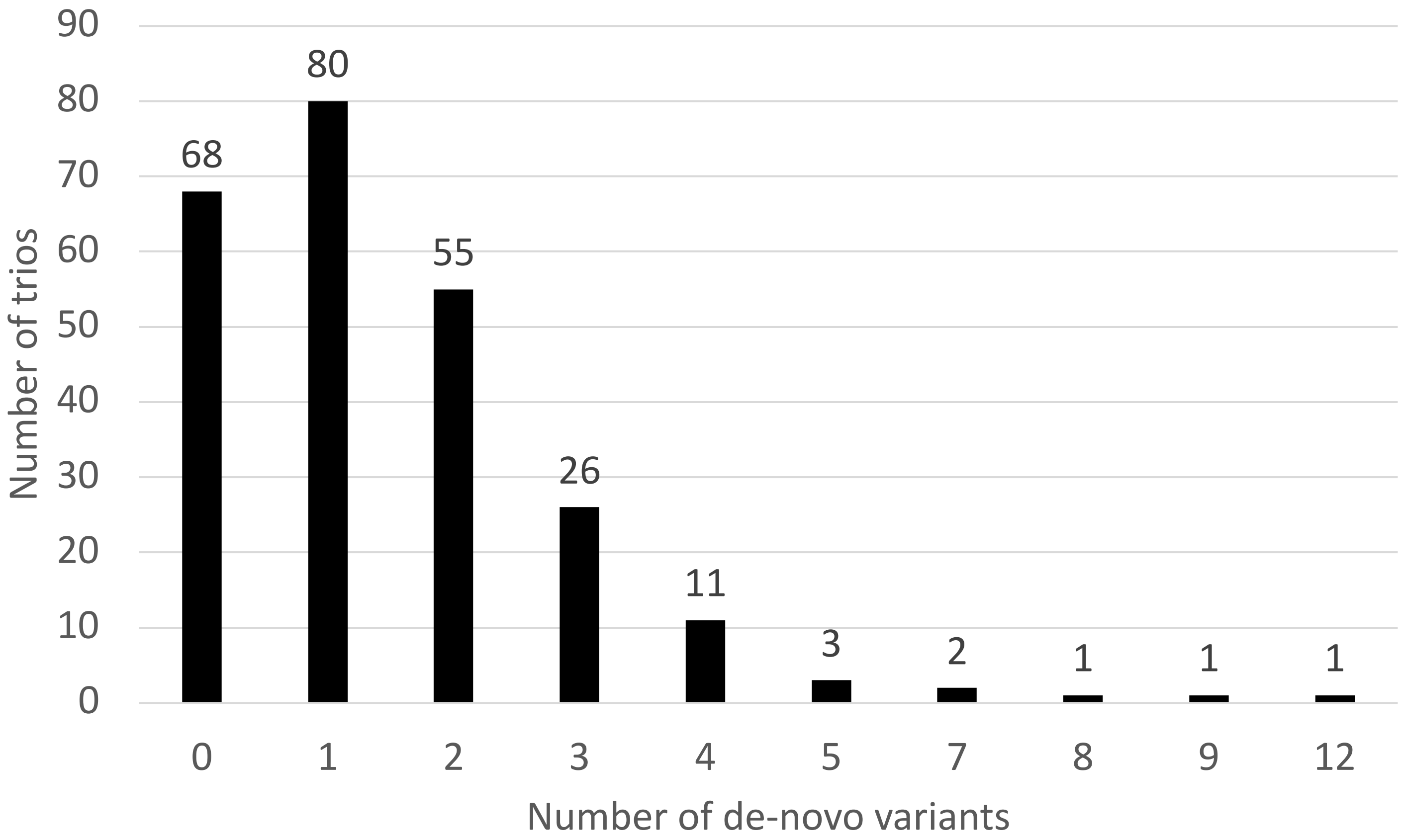

90
80
70
60
50
40
30
20
10
0
Number of trios
68
80
55
26
11
3
2
1
1
1
0
1
2
3
4
5
7
8
9
12
Number of de-novo variants

### Figure S2

**a**

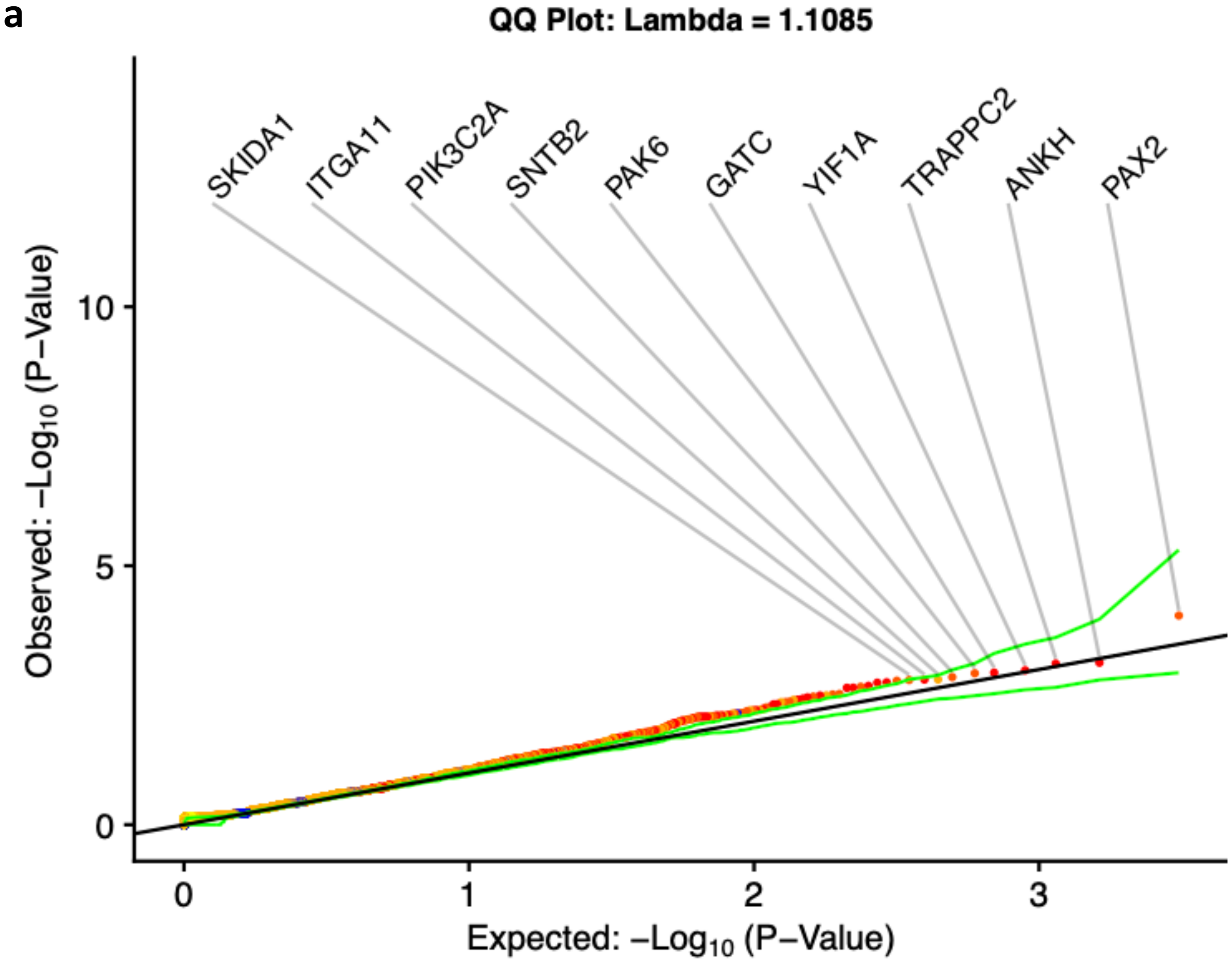

**b**

| Gene | P-value | OR | Case w/wo | Ctrl w/wo |
|---|---|---|---|---|
| *PAX2* | $9.15 * 10^{-5}$ | 11.41 | 7/1735 | 11/22247 |
| *ANKH* | $7.44 * 10^{-4}$ | 22.34 | 4/1738 | 4/22254 |
| *TRAPPC2* | $7.75 * 10^{-4}$ | Inf | 2/1740 | 0/22258 |
| *YIF1A* | $1.04 * 10^{-4}$ | 26.90 | 4/1738 | 2/22256 |
| *GATC* | $1.16 * 10^{-3}$ | Inf | 2/1740 | 0/22258 |
| *PAK6* | $1.19 * 10^{-3}$ | 11.94 | 5/1737 | 5/22253 |
| *SNTB2* | $1.41 * 10^{-3}$ | 12.26 | 5/1737 | 4/22254 |
| *PIK3C2A* | $1.57 * 10^{-3}$ | 4.56 | 9/1733 | 17/22241 |
| *ITGA11* | $1.58 * 10^{-3}$ | 36.99 | 3/1739 | 2/22256 |
| *SKIDA1* | $1.60 * 10^{-3}$ | 9.77 | 5/1737 | 9/22249 |

### Figure S3

**a**

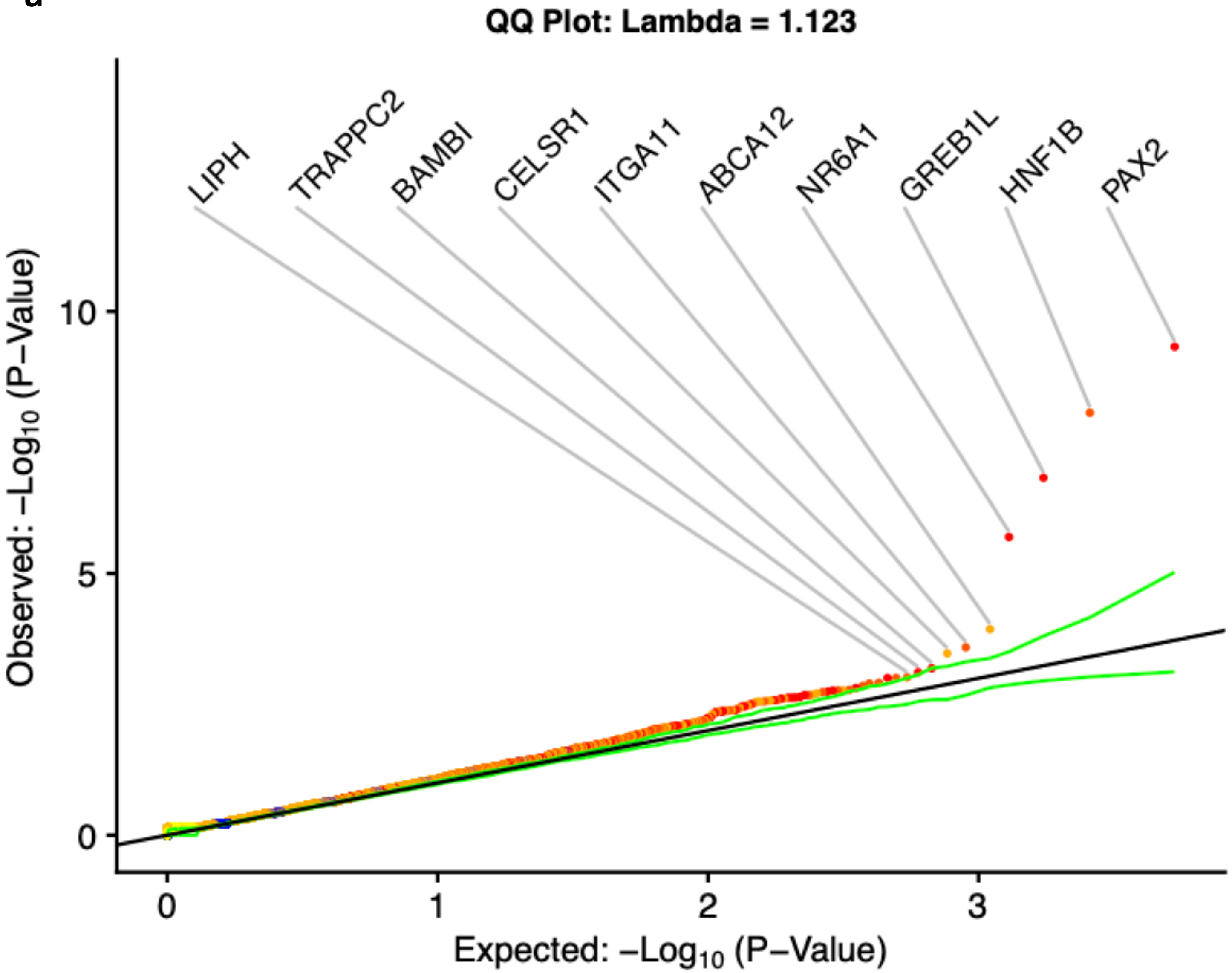

**b**

| Gene | P-value | OR | Case w/wo | Ctrl w/wo |
|---|---|---|---|---|
| *PAX2* | $4.71 * 10^{-10}$ | 19.21 | 14/1728 | 14/22244 |
| *HNF1B* | $8.55 * 10^{-9}$ | 12.57 | 15/1727 | 17/22241 |
| *GREB1L* | $1.50 * 10^{-7}$ | 36.37 | 9/1733 | 4/22254 |
| *NR6A1* | $2.02 * 10^{-6}$ | 16.77 | 9/1733 | 9/22249 |
| *ABCA12* | $1.16 * 10^{-4}$ | 3.37 | 18/1724 | 60/222198 |
| *ITGA11* | $2.59 * 10^{-4}$ | 13.84 | 5/1737 | 12/22246 |
| *CELSR1* | $3.38 * 10^{-4}$ | 3.72 | 14/1728 | 43/22215 |
| *BAMBI* | $6.48 * 10^{-4}$ | 39.67 | 3/1739 | 3/22255 |
| *TRAPPC2* | $7.75 * 10^{-4}$ | Inf | 2/1740 | 0/22258 |
| *LIPH* | $9.66 * 10^{-4}$ | 10.69 | 5/1737 | 9/22249 |

### Figure S4

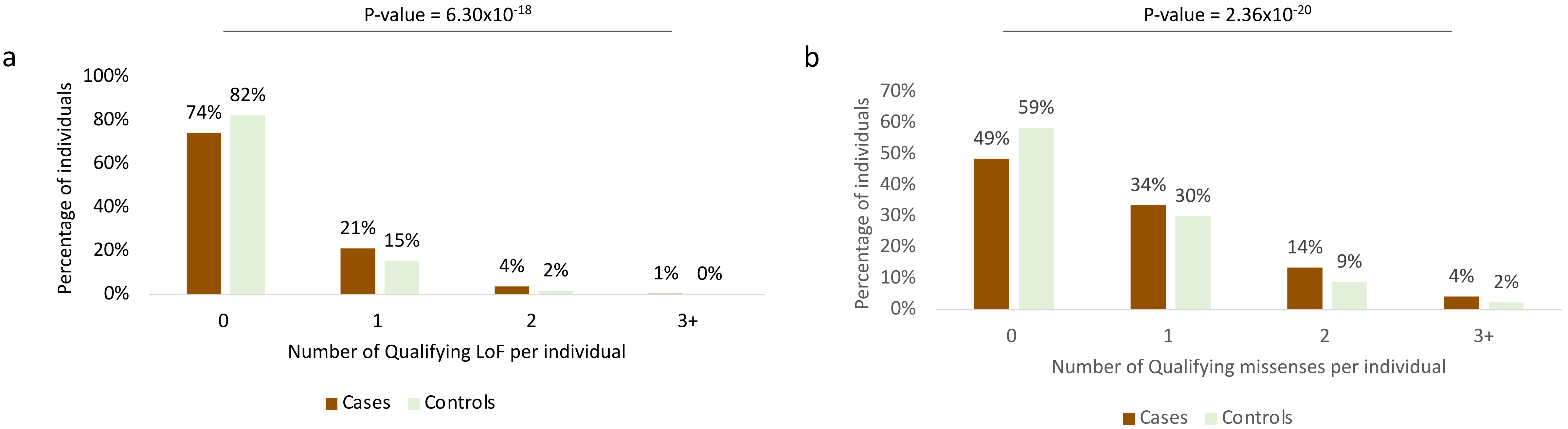
P-value = 6.30x10-18
a
100%
80%
60%
40%
20%
0%
Percentage of individuals
74%
82%
21%
15%
4%
2%
1%
0%
0
1
2
3+
Number of Qualifying LoF per individual
Cases
Controls
P-value = 2.36x10-20
b
70%
60%
50%
40%
30%
20%
10%
0%
Percentage of individuals
49%
59%
34%
30%
14%
9%
4%
2%
0
1
2
3+
Number of Qualifying missenses per individual
Cases
Controls

### Figure S5

a

| Entrez Gene ID | *Symbol* | function | co-expression z-score with NR6A1 | co-expression z-score with ARID3A |
|---|---|---|---|---|
| 1820 | *ARID3A* | AT-rich interaction domain 3A | 4.21 | N/A |
| 2649 | *NR6A1* | nuclear receptor subfamily 6 group A member 1 | N/A | 4.21 |
| 55636 | *CHD7* | chromodomain helicase DNA binding protein 7 | 3.88 | 2.73 |
| 3110 | *MNX1* | motor neuron and pancreas homeobox 1 | N/A | 3.51 |
| 57167 | *SALL4* | spalt like transcription factor 4 | 3.45 | 2.69 |
| 1399 | *CRKL* | CRK like proto-oncogene, adaptor protein | 2.24 | 3.45 |
| 7248 | *TSC1* | TSC complex subunit 1 | 3.25 | N/A |
| 6794 | *STK11* | serine/threonine kinase 11 | N/A | 3.08 |
| 80000 | *GREB1L* | GREB1 like retinoic acid receptor coactivator | 2.99 | N/A |
| 6792 | *CDKL5* | cyclin dependent kinase like 5 | 2.95 | N/A |
| 4038 | *LRP4* | LDL receptor related protein 4 | 2.92 | N/A |
| 4613 | *MYCN* | MYCN proto-oncogene, bHLH transcription factor | N/A | 2.86 |
| 1028 | *CDKN1C* | cyclin dependent kinase inhibitor 1C | N/A | 2.79 |
| 653361 | *NCF1* | neutrophil cytosolic factor 1 | N/A | 2.69 |
| 7546 | *ZIC2* | Zic family member 2 | 2.66 | N/A |
| 23312 | *DMXL2* | Dmx like 2 | 2.62 | N/A |
| 261734 | *NPHP4* | nephrocystin 4 | 2.56 | N/A |
| 139285 | *AMER1* | APC membrane recruitment protein 1 | 2.33 | N/A |
| 203286 | *ANKS6* | ankyrin repeat and sterile alpha motif domain containing 6 | 2.1 | N/A |

b

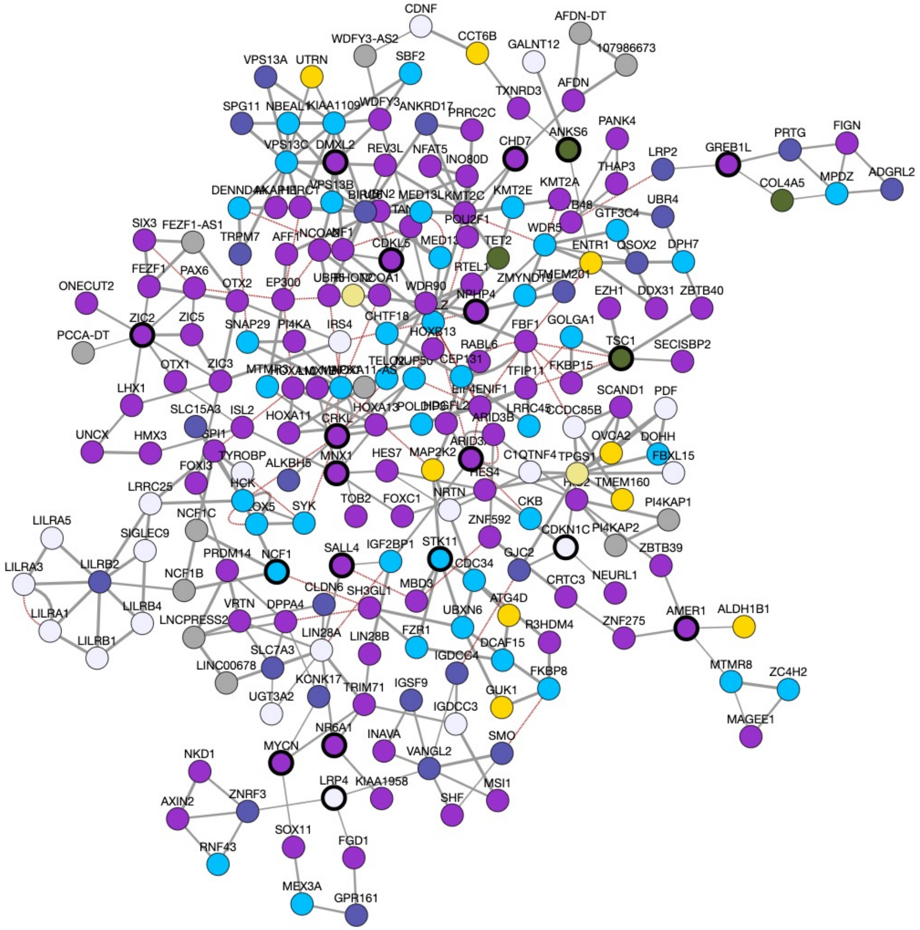

c

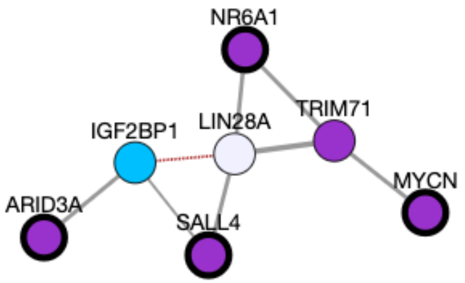

Figure S6

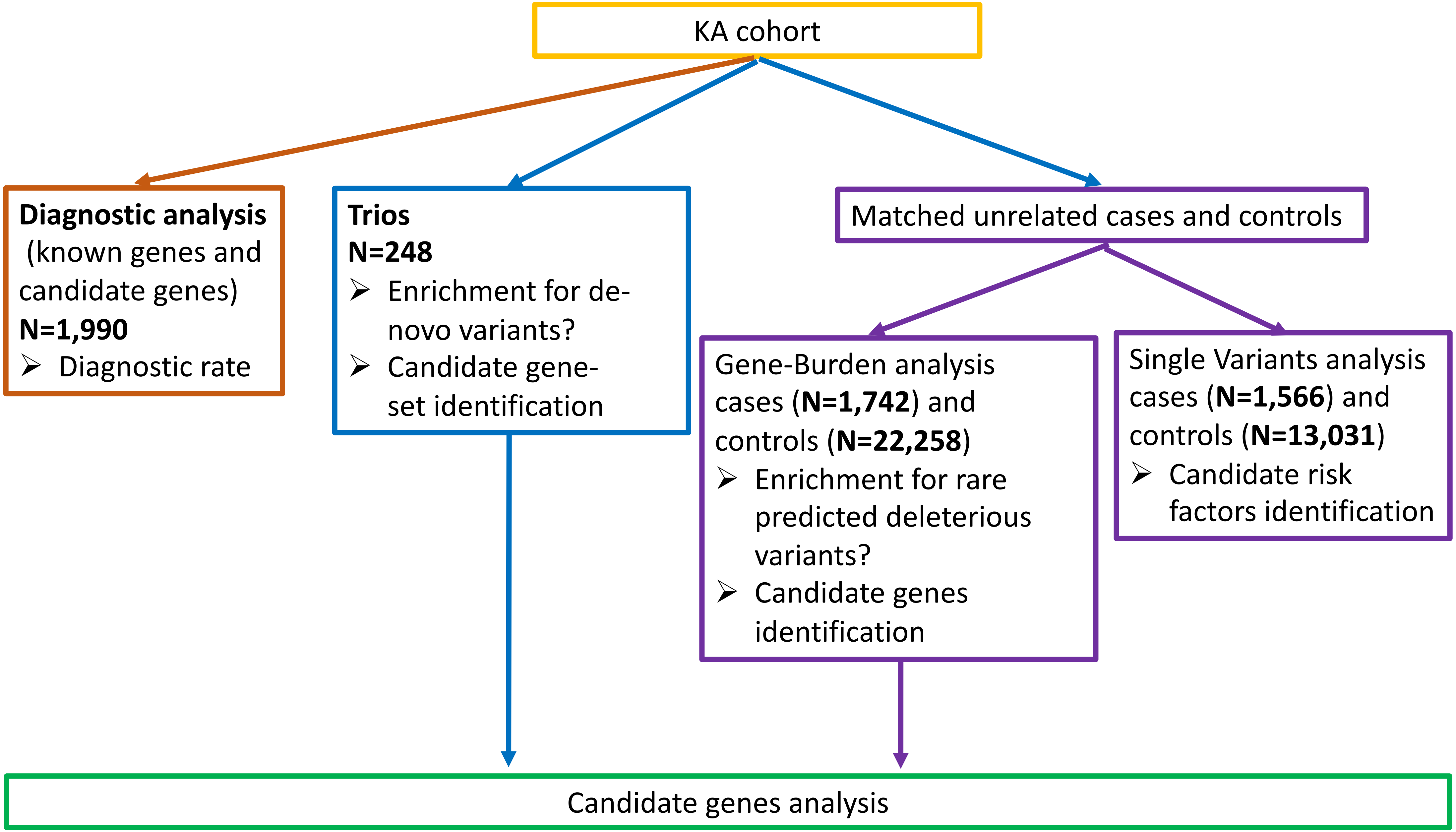

KA cohort
**Diagnostic analysis** (known genes and candidate genes) **N=1,990**
Diagnostic rate
**Trios** **N=248**
Enrichment for de-novo variants?
Candidate gene-set identification
Matched unrelated cases and controls
Gene-Burden analysis cases (**N=1,742**) and controls (**N=22,258**)
Enrichment for rare predicted deleterious variants?
Candidate genes identification
Single Variants analysis cases (**N=1,566**) and controls (**N=13,031**)
Candidate risk factors identification
Candidate genes analysis

**Table S1**: Characteristic of the final set of controls utilized in the case-control analysis

| | | N | Proportion |
|---|---|---|---|
| Total | | 22,258 | |
| Sex | Female | 10,844 | 48.7% |
| | Male | 11,414 | 51.3% |
| Broad phenotype | Control or healthy family members | 9,330 | 42% |
| | Amyotrophic lateral sclerosis | 3,214 | 14% |
| | IgA Nephropathy | 2,387 | 11% |
| | Pulmonary disease | 1,481 | 7% |
| | Ophthalmic disease | 1,155 | 5% |
| | Neuropsychiatric disease [a] | 938 | 4% |
| | Liver and gastrointestinal diseases, | 677 | 3% |
| | Adult Dementia | 763 | 3% |
| | Infectious disease | 344 | 2% |
| | IgA vasculitis | 273 | 1% |
| | Neurological disease [b] | 271 | 1% |
| | Complement 3 glomerulopathy | 154 | 1% |
| | Other [c] | 1271 | 6% |
| Genetic ancestry (PCA based) | Europe | 13,031 | 59% |
| | South America (Latino/Hispanic) | 2,961 | 13% |
| | Africa | 2,715 | 12% |
| | Middle East | 584 | 3% |
| | Asia | 444 | 2% |
| | Admixed | 2,523 | 11% |
| Exome kit | Roche | 13,856 | 62.3% |
| | RocheV2 | 75 | 0.3% |
| | IDTERPv1 [d] | 7,756 | 34.8% |
| | IDTERPv2 | 55 | 0.2% |
| | Agilentv5 | 299 | 1.3% |
| | AgilentV4 | 217 | 1.0% |

[a] Obsessive compulsive disorder, Schizophrenia and others
[b] Ataxia, parkinsonism and others
[c] Hearing loss, Cerebral palsy, hematological diseases, primary immune deficiency and other
[d] Does not include *GREB1L*

**Table S2**: Gene list used for diagnostic analysis (Excel file)

**Table S3**: Diagnostic variants (SNVs, indels and CNVs)- Excel file

**Table S4**: De-novo enrichment analysis in 248 trios

| Model | Variant type | # genes | Observed | | Expected | | Enrichment | p-value | FDR q-value | |
|---|---|---|---|---|---|---|---|---|---|---|
| | | | n | Rate | n | Rate | | | | |
| Genome-wide | Total | 18,931 | 369 | 1.49 | 275.3 | 1.11 | 1.34 | 4.46 $\times10^{-8}$ | 8.96 $\times10^{-7}$ | *** |
| | Synonymous | 18,931 | 78 | 0.31 | 81.6 | 0.33 | 0.95 | 0.67 | 0.77 | |
| | Missenses | 18,931 | 246 | 0.99 | 180.5 | 0.73 | 1.36 | 2.18 $\times10^{-6}$ | 1.54 $\times10^{-5}$ | ** |
| | LoF | 18,931 | 45 | 0.18 | 24.3 | 0.10 | 1.85 | 1.14 $\times10^{-4}$ | 3.42 $\times10^{-4}$ | * |
| | LoF and Missenses | 18,931 | 291 | 1.17 | 204.9 | 0.83 | 1.42 | 8.76 $\times10^{-9}$ | 3.94 $\times10^{-7}$ | *** |
| Constrained genes | Total | 3039 | 109 | 0.44 | 67.7 | 0.27 | 1.61 | 2.39 $\times10^{-6}$ | 1.02 $\times10^{-5}$ | ** |
| | Synonymous | 500 | 3 | 0.01 | 5 | 0.02 | 0.60 | 0.87 | 0.88 | |
| | Missenses | 951 | 30 | 0.12 | 16.6 | 0.07 | 1.8 | 2.01 $\times10^{-3}$ | 6.03 $\times10^{-3}$ | |
| | LoF | 2850 | 18 | 0.07 | 5.7 | 0.02 | 3.14 | 3.25 $\times10^{-5}$ | 9.75 $\times10^{-4}$ | * |
| | LoF and Missenses | 3039 | 93 | 0.38 | 50.6 | 0.20 | 1.84 | 5.97 $\times10^{-8}$ | 1.01 $\times10^{-6}$ | *** |
| NPCs and constrained genes | Total | 662 | 30 | 0.12 | 10.7 | 0.04 | 2.8 | 1.04 $\times10^{-6}$ | 5.89 $\times10^{-6}$ | *** |
| | Synonymous | 662 | 3 | 0.01 | 3 | 0.01 | 1.00 | 0.58 | 0.76 | |
| | Missenses | 237 | 11 | 0.04 | 2.9 | 0.01 | 3.78 | 2.26 $\times10^{-4}$ | 1.35 $\times10^{-3}$ | |
| | LoF | 631 | 8 | 0.03 | 1 | 4.03 $\times10^{-3}$ | 7.64 | 1.42 $\times10^{-5}$ | 8.52 $\times10^{-5}$ | ** |
| | LoF and Missenses | 662 | 27 | 0.11 | 8.2 | 0.03 | 3.3 | 1.63 $\times10^{-7}$ | 1.39 $\times10^{-6}$ | *** |
| E15.5 and constrained genes | Total | 491 | 18 | 0.07 | 7.4 | 0.03 | 2.42 | 7.25 $\times10^{-4}$ | 2.05 $\times10^{-3}$ | |
| | Synonymous | 491 | 1 | 4.03 $\times10^{-3}$ | 2.1 | 8.47 $\times10^{-3}$ | 0.48 | 0.88 | 0.88 | |
| | Missenses | 201 | 7 | 0.03 | 2.4 | 9.68 $\times10^{-3}$ | 2.96 | 0.01 | 0.02 | |
| | LoF | 457 | 6 | 0.02 | 0.7 | 2.82 $\times10^{-3}$ | 8.74 | 8.11 $\times10^{-5}$ | 1.62 $\times10^{-4}$ | * |
| | LoF and Missenses | 491 | 17 | 0.07 | 5.7 | 0.02 | 3.01 | 8.74 $\times10^{-5}$ | 2.97 $\times10^{-4}$ | * |
| Intellectual disability and/or autism spectrum disorder and | Total | 1,043 | 44 | 0.18 | 28.9 | 0.12 | 1.52 | 0.01 | 0.01 | |
| | Synonymous | 1,043 | 6 | 0.02 | 8.5 | 0.03 | 0.71 | 0.85 | 0.88 | |
| | Missenses | 461 | 14 | 0.06 | 9.3 | 0.04 | 1.5 | 0.09 | 0.09 | |
| | LoF | 962 | 10 | 0.04 | 2.4 | 0.01 | 4.11 | 2.23 $\times10^{-4}$ | 3.35 $\times10^{-4}$ | * |
| | LoF and Missenses | 1,043 | 38 | 0.15 | 21.6 | 0.09 | 1.76 | 8.87 $\times10^{-4}$ | 2.15 $\times10^{-3}$ | |

| | | | | | | | | | |
|---|---|---|---|---|---|---|---|---|---|
| constrained genes | | | | | | | | | |
| CHD and constrained genes | Total | 166 | 9 | 0.04 | 5 | 0.02 | 1.8 | 0.07 | 0.09 |
| | Synonymous | 166 | 0 | 0 | 2.2 | 8.87 x10-3 | 0 | 1 | 1 |
| | Missenses | 67 | 5 | 0.02 | 1.5 | 0.01 | 3.24 | 0.02 | 0.03 |
| | LoF | 153 | 1 | 4.03 x10-3 | 0.4 | 1.61 x10-3 | 2.43 | 0.34 | 0.34 |
| | LoF and Missenses | 166 | 9 | 0.04 | 3.7 | 0.02 | 2.42 | 0.01 | 0.03 |
| Immune and constrained genes | Total | 217 | 9 | 0.04 | 4.3 | 0.02 | 2.09 | 0.03 | 0.05 |
| | Synonymous | 217 | 1 | 4.03 $x10^{-3}$ | 1.2 | 4.84 $x10^{-3}$ | 0.81 | 0.71 | 0.86 |
| | Missenses | 96 | 5 | 0.02 | 1.5 | 6.05 $x10^{-3}$ | 3.44 | 0.02 | 0.03 |
| | LoF | 195 | 2 | 8.06 $x10^{-3}$ | 0.4 | 1.61 $x10^{-3}$ | 5.18 | 0.06 | 0.07 |
| | LoF and Missenses | 217 | 8 | 0.03 | 3.3 | 0.01 | 2.45 | 0.02 | 0.03 |

LoF: Loss of Function variants (stop-gained, stop-lost, start-lost, splice-site and frameshift variants)

*** FDR q-value<$10^{-5}$; ** FDR q-value<$10^{-4}$; * FDR q-value<$10^{-3}$

Constrained: for LoF: pLi>0.9 and oe_lof_up_<0.35, for missenses: misZ>3.09, for LoF and missenses, pLi>0.9 and oe_lof_up_<0.35 and/or misZ>3.09

#### Table S5: Gene-set analysis using case-control cohort

| Gene-set | Model | p-value | FDR q-value | OR | Qualified Case | Number of genes with qualifying variants | Qualified Ctrl | Number of genes with qualifying variants |
|---|---|---|---|---|---|---|---|---|
| Constrained Genes | Syn [a] | 0.06 | 0.09 | 1.11 | 818 | 366 | 8,963 | 467 |
| | LoF | $2.37 \times 10^{-10}$ | $4.03 \times 10^{-9}$ | 1.48 | 447 | 424 | 3913 | 1756 |
| | Mis. | $1.01 \times 10^{-5}$ | $4.29 \times 10^{-5}$ | 1.26 | 896 | 580 | 9,224 | 910 |
| Human NPCs (18 weeks) | Syn | 0.98 | 0.98 | 1.00 | 901 | 500 | 10,384 | 656 |
| | LoF | $3.97 \times 10^{-5}$ | $1.12 \times 10^{-4}$ | 1.70 | 90 | 78 | 621 | 302 |
| | Mis. | $1.11 \times 10^{-3}$ | $2.70 \times 10^{-3}$ | 1.32 | 196 | 120 | 1,672 | 220 |
| Mouse embryonic kidney (E15.5) | Syn | 0.14 | 0.18 | 1.08 | 722 | 379 | 8,034 | 490 |
| | LoF | $3.80 \times 10^{-5}$ | $1.12 \times 10^{-4}$ | 1.86 | 68 | 48 | 414 | 211 |
| | Mis. | 0.16 | 0.19 | 1.15 | 149 | 96 | 1,489 | 193 |
| Intellectual disability or Autism Spectrum Disease | Syn | 0.01 | 0.017 | 1.20 | 1,451 | 887 | 16,704 | 1,028 |
| | LoF | $1.10 \times 10^{-7}$ | $9.18 \times 10^{-7}$ | 1.63 | 177 | 146 | 1,377 | 589 |
| | Mis. | $4.58 \times 10^{-7}$ | $2.06 \times 10^{-6}$ | 1.32 | 636 | 308 | 6,068 | 440 |
| Congenital Heart Disease | Syn | 0.6 | 0.6 | 1.03 | 489 | 144 | 5,617 | 167 |
| | LoF | $2.25 \times 10^{-2}$ | 0.07 | 1.60 | 34 | 25 | 242 | 91 |
| | Mis. | $4.46 \times 10^{-2}$ | 0.07 | 1.23 | 127 | 48 | 1,175 | 68 |
| Immune | Syn | 0.07 | 0.10 | 1.12 | 456 | 163 | 4,972 | 214 |
| | LoF | 0.24 | 0.27 | 1.26 | 32 | 28 | 276 | 105 |
| | Mis. | 0.30 | 0.32 | 1.11 | 117 | 52 | 1,174 | 90 |

LoF: Loss of Function variants (stop-gain, splice-site and indel variants); Syn: synonymous variants; Mis: missenses; OR: odds-ratio

[a] The synonymous model for the "constrained" gene set is a set of random 500 genes picked for both DNV and case-control analysis

**Table S6: P**articipants' distribution by number of rare variants

| **Number of variants per individual** | **Loss-of-function variants in genes constrained against LoF [a]** | | **Qualified missense variants in genes constrained against missenses [b]** | | **LoF or qualified missense variants in constrained genes [c]** | |
|---|---|---|---|---|---|---|
| | Cases | Controls | Cases | Controls | Cases | Controls |
| 0 | 74.3% (n=1295) | 82.4% (n=18345) | 48.6% (n=846) | 58.6% (n=13034) | 12.28%(n=214) | 22.48 (n=5003) |
| 1 | 21.3% (n=371) | 15.5% (n=3441) | 33.6% (n=586) | 30.1% (n=6703) | 25.55% (n=445) | 30.14% (n=6708) |
| 2 | 3.85% (n=67) | 1.85% (n=412) | 13.5% (n=236) | 9% (n=2003) | 25.72% (n=448) | 23.07% (n=5134) |
| 3+ | 0.52% (n=9) | 0.27% (n=60) | 4.25% (n=74) | 2.33% (n=518) | 36.45% (n=635) | 24.32% (n=5413) |
| **Pearson's Chi-squared test p-value** | $6.30 \times 10^{-18}$ | | $2.36 \times 10^{-20}$ | | $1.73 \times 10^{-40}$ | |

Constrained thresholds: [a] for LoF: pLi>0.9 and oe_lof_up_<0.35, [b] for missenses: misZ>3.09, [c] for LoF pLi>0.9 and oe_lof_up_<0.35 and for missenses misZ>3.09

**Table S7**: Novel candidate genes- Excel file

**Table S8: Low frequency variants in genes known to be associated with KA**

| Group | *Gene* | Number of variants with p-value <0.05 and OR>0 | Top QV rsID | p-value | OR | Protein change | REVEL | Alpha missense | GnomAD AF | # Cases | Cases AF | # Ctrls | Controls AF |
|---|---|---|---|---|---|---|---|---|---|---|---|---|---|
| Genes known to be associated with AD KA | *DSTYK* | 1 | rs201091809 | $4.27 \times 10^{-4}$ | 2.39 | p.? | - | - | $3.29 \times 10^{-4}$ | 5 | $1.28 \times 10^{-3}$ | 13 | $4.99 \times 10^{-4}$ |
| | *KMT2D* | 3 | rs112170602 | $5.10 \times 10^{-5}$ | 1.52 | p.Pro3665Ala | 0.21 | 0.059 | $1.18 \times 10^{-3}$ | 10 | $3.19 \times 10^{-3}$ | 61 | $2.26 \times 10^{-3}$ |
| Genes known to be associated with AR KA | *PKHD1* | 3 | NA | $2.89 \times 10^{-4}$ | 2.84 | p.Thr3628Ile | 0.17 | 0.105 | $4.57 \times 10^{-5}$ | 8 | $2.55 \times 10^{-3}$ | 2 | $7.68 \times 10^{-5}$ |
| | *SDCCAG8* | 1 | rs201869920 | $5.97 \times 10^{-4}$ | 1.94 | p.Leu115Val | 0.16 | 0.093 | $4.66 \times 10^{-4}$ | 6 | $1.92 \times 10^{-3}$ | 14 | $4.99 \times 10^{-4}$ |

OR>1.5, p-value<$10^{-3}$

**Table S9**: Sequencing and array platforms used in the KA cases

| | | N | Proportion | Analyzed with XHMM (N) | Proportion | Analyzed with SNP arrays (N) | Proportion |
|---|---|---|---|---|---|---|---|
| Total | | 1,990 | | 1878 | | 1342 | |
| Exome kit | IDTERPv1 | 1,474 | 74% | 1385 | 74.10% | 1117 | 83.23% |
| | IDTERPv2 | 254 | 13% | 253 | 13.54% | 99 | 7.38% |
| | Roche | 36 | 2% | 35 | 1.87% | 26 | 1.94% |
| | RocheV2 | 98 | 5% | 94 | 5.03% | 79 | 5.89% |
| | AgilentV4 | 30 | 2% | 11 | 0.59% | 21 | 1.56% |
| | AgilentV5 | 97 | 5% | 91 | 4.87% | 0 | 0% |
| | 50Mb | 1 | <1% | 0 | 0% | 0 | 0% |
| Array platforms | | | | | | | |
| Array platforms | Illumina MegaEx | 588 | 29.55% | 576 | 30.82% | 588 | 43.82% |
| | Illumina MEGA V1.0 | 348 | 17.49% | 322 | 17.23% | 348 | 25.93% |
| | Illumina Omni 1M | 108 | 5.43% | 95 | 5.08% | 108 | 8.05% |
| | Illumina 610-Quad | 93 | 4.67% | 83 | 4.44% | 93 | 6.93% |
| | Illumina MEGA V1.1 | 70 | 3.52% | 63 | 3.37% | 70 | 5.22% |
| | Illumina Omni 2.5M | 61 | 3.07% | 61 | 3.26% | 61 | 4.55% |
| | Illumina OmniExpress | 58 | 2.91% | 54 | 2.89% | 58 | 4.32% |
| | Illumina 660W | 16 | 0.80% | 12 | 0.64% | 16 | 1.19% |

**Table S10**: Grouping of cases and controls based on genetic ancestry

| cluster | Total | case | control | ratio |
|---|---|---|---|---|
| 0 | 6262 | 130 | 6132 | 0.021 |
| 1 | 4304 | 829 | 3475 | 0.24 |
| 2 | 2766 | 50 | 2716 | 0.018 |
| 3 | 2689 | 337 | 2352 | 0.14 |
| 4 | 2035 | 34 | 2001 | 0.017 |
| 5 | 1979 | 21 | 1958 | 0.011 |
| 6 | 1504 | 267 | 1237 | 0.22 |
| 7 | 1133 | 24 | 1109 | 0.022 |
| 8 | 838 | 29 | 809 | 0.036 |
| 9 | 490 | 21 | 469 | 0.045 |
